# The Florida Pancreas Collaborative Next-Generation Biobank: State-wide Infrastructure to Reduce Disparities and Improve Survival for a Racially and Ethnically Diverse Cohort of Patients with Pancreatic Cancer

**DOI:** 10.1101/2020.10.10.20209247

**Authors:** Jennifer B. Permuth, Kaleena B. Dezsi, Shraddha Vyas, Karla N. Ali, Toni L. Basinski, Ovie A. Utuama, Jason W. Denbo, Jason Klapman, Aamir Dam, Estrella Carballido, DaeWon Kim, Jose M. Pimiento, Benjamin D. Powers, Jung W. Choi, Dung-Tsa Chen, Jamie K. Teer, Francisca Beato, Alina Ward, Elena M. Cortizas, Suzanne Y. Whisner, Iverson E. Williams, Andrea N. Riner, Kenneth Tardif, Vic Velanovich, Andreas Karachristos, Wade G. Douglas, Adrian Legaspi, Bassan Allan, Kenneth Meredith, Manual A. Molina-Vega, Philip Bao, Jamii St. Julien, Kevin L. Huguet, B. Lee Green, Folakemi T. Odedina, Nagi B. Kumar, Vani N. Simmons, Thomas J. George, Mokenge Malafa, Pamela Hodul, Juan P. Arnoletti, Ziad T. Awad, Debashish Bose, Kun Jiang, Barbara A. Centeno, Clement K. Gwede, Sarah M. Judge, Andrew R. Judge, Daniel Jeong, Mark Bloomston, Nipun B. Merchant, Jason B. Fleming, Jose G. Trevino, on behalf of the Florida Pancreas Collaborative

**Affiliations:** Department of Cancer Epidemiology, H. Lee Moffitt Cancer Center & Research Institute, Tampa, FL; Department of Gastrointestinal Oncology, H. Lee Moffitt Cancer Center & Research Institute, Tampa, FL; Department of Diagnostic Imaging, H. Lee Moffitt Cancer Center & Research Institute, Tampa, FL; Department of Biostatistics and Bioinformatics, H. Lee Moffitt Cancer Center & Research Institute, Tampa, FL; Regional Cancer Center, Fort Myers, FL; University of Miami Miller School of Medicine, Miami, FL; Cancer Institute, Advent Health Orlando, Orlando, FL; Department of Surgery, University of Florida, Gainesville, FL; Department of Surgery, St. Anthony’s Hospital, St. Petersburg, FL; University of South Florida/Tampa General Hospital, Tampa, FL; Florida State University College of Medicine, Department of Clinical Scienes, Division of Surgery, Tallahassee Memorial Healthcare, Tallahassee, FL; Center for Advanced Surgical Oncology at Palmetto General Hospital, Tenet Healthcare Palmetto General, Hialeah, FL; Sarasota Memorial Hospital, Sarasota, FL; Lakeland Regional Health, Lakeland, FL; Department of Surgical Oncology, Mount Sinai Medical Center, Miami Beach, FL; Department of Health Outcomes and Behavior, H. Lee Moffitt Cancer Center & Research Institute, Tampa, FL; University of Florida, Gainesville, FL; Department of Medicine; Division of Oncology, University of Florida, Gainesville, FL; Surgical Oncology, Advent Health Orlando, Orlando, FL; Surgery, University of Florida - Jacksonville, Jacksonville, FL; Surgical Oncology, University of Florida-Orlando, Orlando, FL; Department of Anatomic Pathology, H. Lee Moffitt Cancer Center & Research Institute, Tampa, FL; Department of Physical Therapy, University of Florida, Gainesville, FL; Lee Memorial Hospital, Fort Myers, FL; Department of Surgical Oncology, University of Miami Miller School of Medicine, Miami, FL

**Author notes:** **Corresponding Author:** Jennifer B. Permuth, PhD, 12902 USF Magnolia Dr., Tampa, FL 33612.

**Keywords:** Biorepository, underserved populations, cancer disparities, prospective cohort, pancreatic cancer

## Abstract

**Background:** Well-annotated, high-quality biorepositories provide a valuable platform to support translational research and discovery. However, most biorepositories have poor representation of minority groups, limiting the ability to address cancer health disparities and improve disease outcomes. This report describes the establishment of the Florida Pancreas Collaborative (FPC), the first state-wide prospective longitudinal cohort study and biorepository specifically designed to address the higher burden of pancreatic cancer (PaCa) in African Americans (AA) compared to Non-Hispanic Whites (NHW) and Hispanic/Latinx (H/L).

**Methods:** We describe rationale for establishing the FPC and provide an overview of key stakeholders; study eligibility and design; ascertainment and recruitment strategies; and standard operating procedures (SOPs) developed to collect, process, store, and transfer biospecimens, medical images, and data. We also describe the customized cloud-based, secure data management platform built to facilitate recruitment, track study-related workflow, house data, and perform queries. We also present progress to date regarding recruitment and biobanking.

**Results:** The FPC consists of multidisciplinary teams from fifteen Florida medical institutions. From March 2019 through August 2020, 350 patients were assessed for study eligibility, 323 met inclusion/exclusion criteria, and 305 (94%) enrolled, including 228 NHW, 30 AA, and 47 H/L, with 94%, 100%, and 94% participation rates, respectively. A high percentage of participants have donated blood (87%), pancreatic tumor tissue (41%), computed tomography scans (76%), and baseline questionnaire data (62%).

**Conclusions:** This biorepository addresses a critical gap in PaCa research with the potential to advance basic, clinical, population-based, and translational studies intended to minimize disparities, increase quality of life, and reduce PaCa-related morbidity and mortality.

**Impact:** This multi-institutional infrastructure can serve as a prototype for development of similar resources across the country and disease sites.

## Introduction

Pancreatic cancer (PaCa) is the deadliest solid malignancy in the United States (US), with a five-year relative survival rate of 10%^1^. An estimated 57,600 Americans will be diagnosed with PaCa and 47,050 will die from the disease in 2020^1^. Surgical resection offers the only chance for long-term survival, but only 15-20% of cases are resectable at diagnosis. Due to the lack of effective strategies for prevention, early detection, and treatment, PaCa is projected to become the second leading cause of cancer deaths by 2030^2^. Coinciding with the rise in the number of PaCa diagnoses and deaths is a notable health disparity^2-12^, with African Americans (AA)/Blacks having 25-40% and 25-56% higher PaCa incidence and mortality rates, respectively, compared to Non-Hispanic Whites (NHW) and Hispanic/Latinx (H/L) ^3^.

The state of Florida has the third largest population in the US, is home to more than 3.5 million AA^13^ and 6.1 million H/L^14^, and is surpassed only by California in lives lost to PaCa each year^1^. In 2020, 3,570 (7.9%) of the 45,300 cancer-related deaths among Floridians will be due to PaCa and occur mainly in NHW, AA, and H/L^15^. Despite these statistics, PaCa health disparities research is limited among Florida’s diverse population. The Surveillance, Epidemiology and End Results (SEER) Registry captures cancer statistics for nearly one-third of the US population^3^ and reveals higher PaCa incidence and mortality rates for AA adults compared to other groups. Although Florida is not included in the SEER Registry, we used Florida Cancer Data System (FCDS) Registry data^16^ to estimate age-adjusted PaCa incidence and mortality rates for self-reported AA, NHW, and H/L and found that AA had the highest incidence and mortality rates across genders, mirroring national disparities^17^. Thus, strong rationale exists for studying and addressing disparities in Florida’s large and diverse PaCa population which is representative of the burden across the US.

Most PaCa disparities research has been descriptive, with inequities unexplained by epidemiologic and socioeconomic factors. Modifiable PaCa risk factors which can also contribute to poor outcomes include tobacco exposure, being overweight (body mass index (BMI) ≥25 kg/m^2^) or obese (BMI ≥30 kg/m^2^), and having a history of chronic or new onset diabetes^18-36^. Although early case-control studies^37^ reported excess PaCa incidence among AA to be explained by smoking and diabetes in men and high BMI in women, a more recent analysis showed higher AA PaCa incidence persists after controlling for smoking, diabetes, obesity, and family history^38^. Socioeconomic factors, access barriers, and bias also influence PaCa disparities. Compared to NHW, AA are less likely to: be referred to PaCa specialists, be diagnosed/treated at high-volume hospitals, receive surgery or chemotherapy, be insured, or have high incomes^11,37,39-43^. Moreover, structural racism at individual and institutional levels may be a driver of these racial health inequities^44^. Since interventions to reduce disparities can be more successful when the underlying mechanisms are understood^45^, evaluation of biological factors that contribute to disparities is critical. Obstacles to the discovery of mechanisms of pancreatic carcinogenesis among AA include the lack of biospecimens, clinical data, radiologic images, and preclinical models.

Biorepositories provide a rich platform to study and address cancer health disparities and improve clinical outcomes. High-quality, well-annotated national^46^ and institutional^47^ biorepositories have been developed to better understand the pathophysiology and biological mechanisms of PaCa and related conditions (chronic pancreatitis and diabetes^46^), but addressing health disparities was not a focus in their development and biospecimens from minority groups are scarce in these resources. Our primary objective was to develop a platform for translational PaCa disparities studies by building a robust state-wide ‘next-generation biobank’ containing viable tissues, biofluids, images, and clinical, epidemiologic, and laboratory data, with a racial/ethnically diverse cohort of patients with PaCa and its precursors.

## Methods

### Assembling a Team of Key Stakeholders

#### Participating sites, multidisciplinary expertise, and advisory boards

The Florida Pancreas Collaborative (FPC) was founded in 2015 by investigators (JBP, MPM, JGT, NBM) from the three main academic cancer centers in the state of Florida: Moffitt Cancer Center and Research Institute (Tampa, MCC), the University of Florida Health Cancer Center/University of Florida (Gainesville, UFG), and the Sylvester Comprehensive Cancer Center/University of Miami (Miami, UOM)^48^. Florida Agency for Health Care Administration (AHCA)^49^ inpatient discharge data was used to identify institutions throughout Florida with the highest numbers of AA, H/L, and NHW individuals diagnosed and treated for PaCa. We then used internet queries and our professional network to identify and contact clinicians (primarily surgeons and oncologists) to assess interest in participation in the collaborative. Requirements for participation included having a dedicated site principal investigator (PI), institutional support/backing, and willingness to contribute data, biospecimens, and medical images to a common biorepository using standard operating procedures (SOPs). With infrastructure grant funding from the State of Florida’s James and Esther King Biomedical Research Program in 2018, the FPC expanded to include twelve additional institutions (academic and community cancer centers and private hospitals) including (in alphabetical order): Advent Health Orlando (Orlando, AHO), Jackson Memorial Hospital (Miami, JMH), Lakeland Regional Health Hollis Cancer Center (Lakeland, LRH), Mount Sinai Medical Center (Miami, MSM), Palmetto General Hospital (Hialeah, PGH), Regional Cancer Center (Fort Myers, RCC), Saint Anthony’s Hospital/BayCare (St. Petersburg, STA), Sarasota Memorial Hospital (Sarasota, SMH), Tallahassee Memorial Healthcare (Tallahassee, TMH), the University of Florida Health (Jacksonville, UFJ), Orlando Health University of Florida Health Cancer Center (Orlando, UFO), and the University of South Florida/Tampa General Hospital (Tampa, USF). MCC serves as the lead coordination and management center. A map of participating FPC sites is displayed in **Figure 1**.

**Figure 1.**
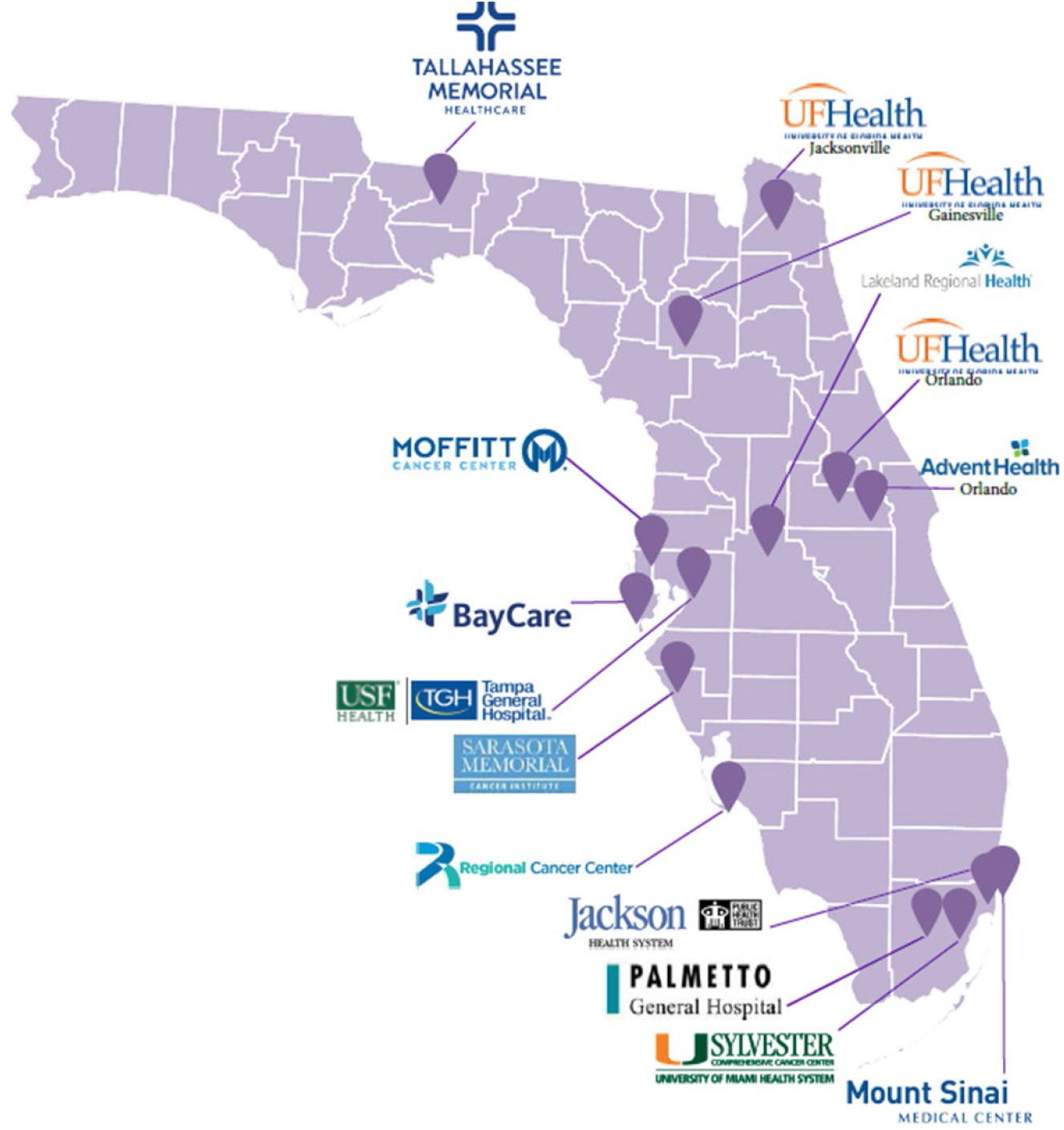
Florida Pancreas Collaborative Study Sites.

Our multidisciplinary, cross-institutional team has expertise in disciplines including molecular epidemiology, surgical oncology, radiology, gastrointestinal endoscopy, medical oncology, radiation oncology, nutrition, genetics, molecular biology, muscle physiology, behavioral science, pathology, biostatistics, and bioinformatics. A scientific advisory board advises, oversees, and evaluates research activities related to the study aims and consists of members from major and regional cancer centers chosen for their research and clinical expertise and strong regional and national leadership in the areas of Diversity/Health Equity Research, PaCa Treatment, Nutrition/Cachexia Research, and Tobacco Cessation. Given that recruiting individuals to participate in biobanks can be challenging^47,50^, we are working with community partners including MCC’s National Cancer Institute (NCI)- funded Tampa Bay Community Cancer Network and the Geographic Management of Cancer Health Disparities Program^143, 192, 193^, the George Edgecomb Society, and local affiliates of the Pancreatic Cancer Action Network to expand our community advisory board which includes PaCa survivors and advocates. Community advisors can be integral to ensuring perspectives and voices of individuals from diverse backgrounds and experiences are heard and to helping set priorities, improve the ethical nature of research, appropriateness of methods, and increase recruitment and retention^51^.

### Overview of the Study Population and Design

#### Study population

To be eligible for participation, an individual of any gender identity must meet the following eligibility criteria: be at least 18 years of age; self-report as NHW, AA, or H/L; present to the gastrointestinal (GI) clinic, surgery, or endoscopy at a participating FPC site with a strong clinical suspicion or diagnosis of a pancreatic tumor based on symptoms, imaging, biopsy, and/or blood-work; have a treatment-naïve pancreatic tumor at the time of enrollment; be able to understand and voluntarily sign the informed consent; and be willing to complete study questionnaire(s) and donate medical images and biospecimens at the time of standard of care procedures after signing the informed consent document. Confirmation of the diagnosis is sought for all cases and is typically confirmed by pathologic review of tissue specimens obtained through routine diagnostic procedures by a participating site pathologist. In some circumstances, we will be relying on cancer registry data, the electronic medical record (EMR), or report on a death certificate. To increase the breadth of cases for inclusion, we allow enrollment of patients with operable or inoperable exocrine pancreatic tumors (primarily pancreatic ductal adenocarcinomas or PDAC), endocrine pancreatic tumors (such as pancreatic neuroendocrine tumors (PNET)), and patients with pre-malignant cystic lesions known as intraductal papillary mucinous neoplasms (IPMN) and mucinous cystic neoplasms (MCN).

#### Study Design

This three-year research infrastructure grant uses a prospective longitudinal multi-institutional cohort study design. The project began administratively in May of 2018, and a 10-month run-in period was essential for infrastructure building. With input from stakeholders, substantive accomplishments during the run-in period included: meetings with stakeholders to refine the scope of work; development of a study logo, recruitment materials, and study web-site; finalizing a uniform study protocol, informed consent document, study questionnaires, and data collection instruments/case report forms (CRF); translation of study materials into Spanish; obtaining regulatory approval through the single Institutional Review Board (sIRB; Advarra), development of SOPs for data, medical image, and biospecimen collection, processing, storage, and transfer; building a customized cloud-based centralized platform for data collection, management, and workflow; and hiring and training staff. The resources collected at each time-point (baseline, 6 months, and 12 months) are summarized in **Table 1**.

**Table 1.** Summary of Florida Pancreas Collaborative biobank contents and time-points for collection.

| Biobank contents | Time-point |  |  |
| --- | --- | --- | --- |
|  | Baseline | 6 months | 12 months |
| Health Screen | X | X | X |
| Study Questionnaire | X | X | X |
| Clinical, laboratory, imaging, and pathologic data abstracted from the medical record and/or requested from the Florida Cancer Data System | X | X | X |
| Blood processed for plasma and serum (and DNA for ancestry analysis in the future) | X | X | X |
| Tissue from surgery (or biopsy): pancreas tumor (PT), normal pancreas (NP), adipose-subcutaneous (AD-S), adipose-omental (AD-O), and muscle (MU) | X | a | a |
| Computed tomography (CT) images | X | X | X |

<sup>a</sup> Tissue to be collected for research if a procedure is being performed as part of clinical care.

#### Ascertainment and Recruitment Strategies

Engagement of the entire clinical research team is critical to successfully building a biobank. Each participating site has a lead research coordinator who works closely with the site PI and the program manager at the coordinating site. This coordinator is integrated into each clinic’s workflow and is responsible for screening daily clinic and procedure logs to identify individuals to approach regarding participation. To aid in recruitment, we developed a study-specific flyer, informational brochure, and public-facing web-site (www.floridapancreascollaborative.org) with a members-only portal accessible via secure sign-in by participating site staff. Additionally, based on data published by our team and others supporting incentives as motivating factors in increasing study participation^52-55^, all participants receive compensation (in the form of gift card(s)) to Amazon or Walmart) as a token of appreciation for their time and effort upon completion of baseline ($10) and follow-up questionnaires ($5 each).

#### Overview of Study Workflow and Data Management/Tracking

A customized cloud-based data management/ engagement platform was built in partnership with DatStat, Inc (Seattle, Washington). This platform helps each site efficiently perform the workflow summarized in **Supplementary Figure 1** including assignment of unique identification (ID) numbers, assessment of eligibility, obtaining informed consent electronically, administering questionnaires, and tracking biospecimens. Detailed views of the study platform and select components are displayed in **Supplementary Figures 2 and 3**. The platform also manages and stores study-related data and allows queries to be performed. Individual study sites may only access information pertaining to their own study participants while the coordinating site has regulatory approval to access and analyze data across all sites.

### Standard operating procedures (SOPs)

#### Data collection procedures and instruments

At each timepoint, the research coordinator records the participant’s height and weight from the medical chart, measures their waist and hip circumference using standard procedures^56^, and administers a 3-page health screen through the DatStat platform **(Table 1)**. The health screen evaluates the presence of conditions prevalent among patients with PaCa: cancer cachexia, a progressive and debilitating muscle-wasting syndrome characterized by unintentional weight loss, muscle atrophy, fatigue, and limited tolerance of chemotherapy^57,58^; depression and distress^59-62^; and past and present smoking history^18-30^ and assesses parameters such as weight change, dietary intake, symptoms, performance status, and emotional, physical, and practical concerns. The health screen comprises three instruments: the abridged version of the Patient-Generated Subjective Global Assessment (aPG-SGA)^63^, a revised version of the Edmonton Symptom Assessment System (ESAS-r), and the Canadian Problem Checklist^64,65^ **(Supplementary Figure 4)**. This screening tool helps our providers to better understand the concerns of participants and automatically ‘flags’ issues via a customized ‘Health Screen’ report. Our intent is to proactively enhance patient care, outcomes, and experiences through education/counseling and referral to other professionals (dieticians, physical therapists/rehabilitation, psychiatrists, psychologists, social workers, speech pathologists) as needed. An example of a Health Screen Report for a hypothetical participant with probable refractory cachexia, depression, and a current smoking history is depicted in **Supplementary Figure 5**. At baseline, performance status is also assessed by the treating provider using Eastern Cooperative Oncology Group (ECOG) guidelines^66^.

Participants complete web-based or teleform questionnaires. As summarized in **Table 2**, the baseline questionnaire solicits core demographic, clinical, and epidemiologic data elements including gender identity, race/ethnicity, presenting systems, age at diagnosis, history of diabetes and other comorbidities, medication use, alcohol history, family history, physical activity, dietary intake, reproductive history for women, and gender-affirming interventions^67^. It also assesses tobacco use and exposure using questions from the NCI-American Association for Cancer Research (AACR) Cancer Patient Tobacco Use Assessment Task Force^68^ and the Fagerstrom Test for Nicotine Dependence (FTND), a highly recognized tool to quickly identify nicotine dependence with 10 questions^69,70^. Based on reports of enhancements in therapeutic efficacy with use of cannabinoids^71^, participants are asked about medical marijuana which is widely available in the state of Florida, a topic important to the funding agency and the Coalition for Medical Marijuana Research and Education. The baseline questionnaire contains several validated instruments to assess lifestyle factors and domains including mental health, lifetime stress^72^, sleep^73^, nutrition, and quality of life. For example, data regarding dietary intake and physical activity are collected using the Dietary Screener Questionnaire (DSQ) developed by the NCI’s Risk Factor Assessment Branch and the International Physical Activity Questionnaire (IPAQ)-Short Form, respectively. Other instruments include the European Organization for Research and Treatment of Cancer (EORTC) QLQ-PAN26, a questionnaire validated in pancreatic diseases^74-76^ that is administered with core QLQ-30. PAN26 contains 26 items on self-reported symptoms and emotional issues specific to PaCa, with a high score (100%) indicative of more severe symptoms and lower quality of life. We also measure dispositional optimism using the Life Orientation Test-Revised (LOT-R) test and systematically evaluate lifetime exposure to acute and chronic stressors using the online Stress and Adversity Inventory (STRAIN) tool^72^. A list of all validated instruments used in the FPC health screen or baseline questionnaire can be found in **Supplementary Table 1**.

**Table 2.**
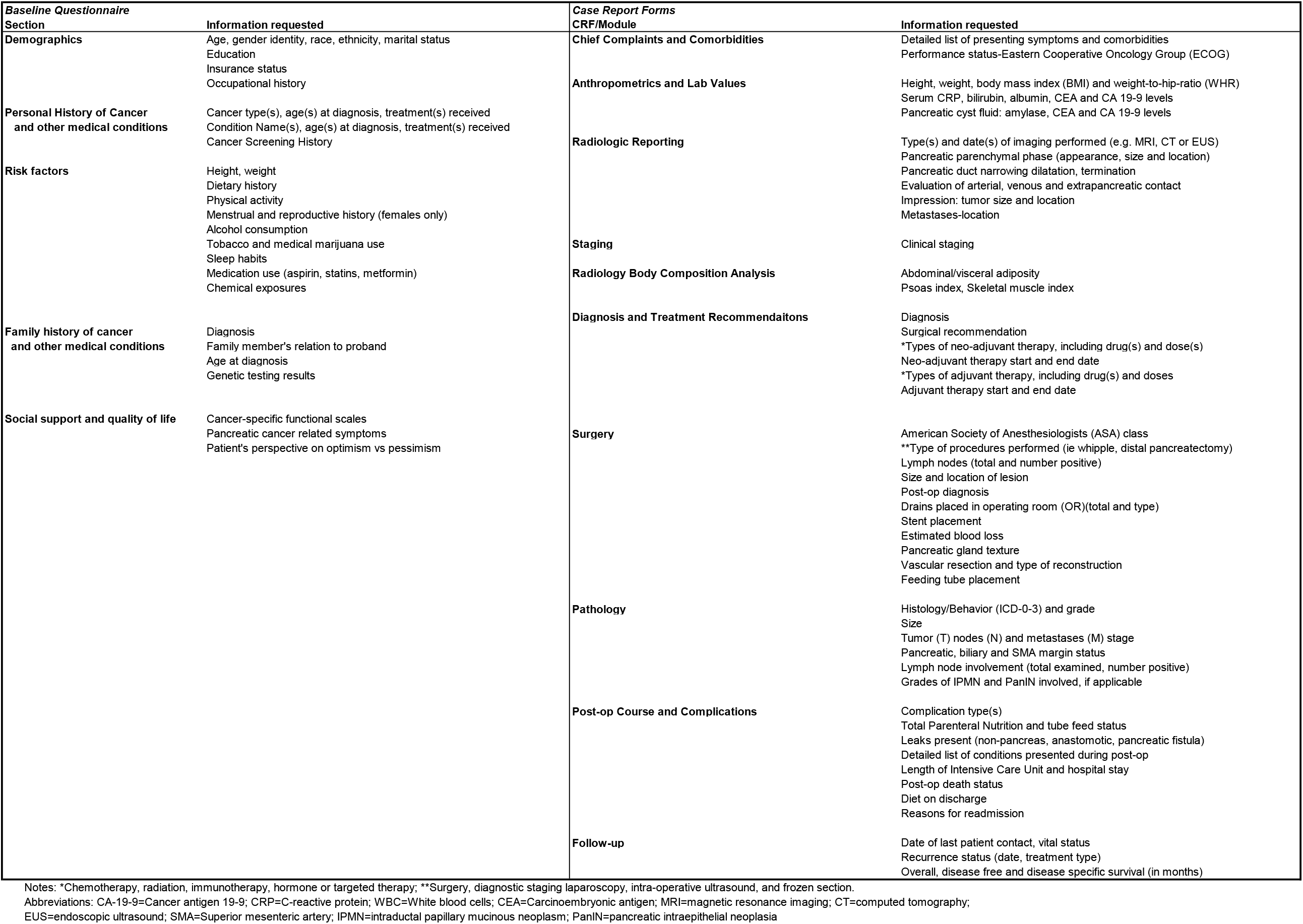
Data elements solicited in the Florida Pancreas Collaborative Baseline Study Questionnaire or Case Report Forms (CRF).

Data from the health screen and self-administered questionnaires is being supplemented by data abstracted from the EMR along with data requested from the FCDS. Supplemental data from the EMR is entered manually into CRFs within the study database by the site research coordinator or by trained staff at the coordinating center. The CRFs, summarized in **Table 2**, capture: 1) presenting signs, symptoms, and comorbidities; 2) pre-treatment work-up details (including anthropometrics and lab values; medical image availability; standardized radiologic reporting criteria^77^; staging information; diagnosis and treatment recommendations; and radiologic metrics of body composition); 3) treatment details (surgery and other modes of chemo-, radiation-, immuno-, hormonal-, or targeted-therapy (neoadjuvant and/or adjuvant); post-operative course and complications; pathology) and 4) follow-up information (date of last contact, vital status, overall and progression free survival). For data management and tracking purposes, blood, tissue, and image collection/ transfer CRFs were also designed.

#### Biospecimen collection, processing, and storage

Differences in biospecimen collection, processing, and storage methods can serve as sources of error in studies of biospecimen discovery, development, and validation, thus SOPs were developed and tested at the coordinating center and modified as needed to ensure compliance at participating centers. All supplies and reagents are provided by the coordinating site. To enable long term storage/preservation, digital bar-code labeled (DBL) cryogenic vials were chosen because of their durability and ability to facilitate accurate data entry and rapid retrieval. The DBL is not linked to the patient name, medical record number, or other forms of institutional patient identification. Freezers at the coordinating site have autonomous continuous temperature monitoring and an alarm system to notify responsible parties of a malfunction. Freezers are on a stable power grid with backup generators and are above ground level to prevent damage by flooding.

##### Blood

Consented participants donate peripheral blood samples at baseline (defined as +/- 30 days of the diagnosis date) and at the time of follow-up (∼6 months and ∼12 months after baseline) in conjunction with routinely-scheduled venipuncture. Using SOPs for blood collection and processing in line with the NCI’s Best Practices^78^, four 10 mL tubes of blood (2 red-top, 2 purple-top EDTA) are collected at baseline and two 10 mL tubes are requested (1 red-top and 1 EDTA) at follow-up visits. The date and time the samples are drawn, processed, and stored, and other details such as the visual assessment of hemolysis are recorded. EDTA tubes are slowly inverted 8 to 10 times to ensure mixing of the sample and the anti-coagulant liquid inside the tube and then transferred to the institution’s laboratory for processing 30 minutes to 2 hours after collection. EDTA tubes are processed first. For samples collected at baseline, 1 mL of whole blood (from each EDTA tube) is aliquoted into designated cryovials. To process for plasma, the remaining blood is centrifuged at 1300g/RT/10min and harvested 12 cryovials with 0.5 mL each (**Figure 2a**). The red-topped tubes are processed for serum after allowing 30 minutes for clotting. Samples are centrifuged @1300g/RT/10min. At baseline, 1 mL serum is aliquoted into each of 6 cryovials and 0.5 mL each into 8 cryovials (**Figure 2b**). For follow-up, cryovial specifics are shown in **Figures 2a and 2b**. All samples are stored in cryoboxes at −80°C until transferred to MCC.

**Figure 2.**
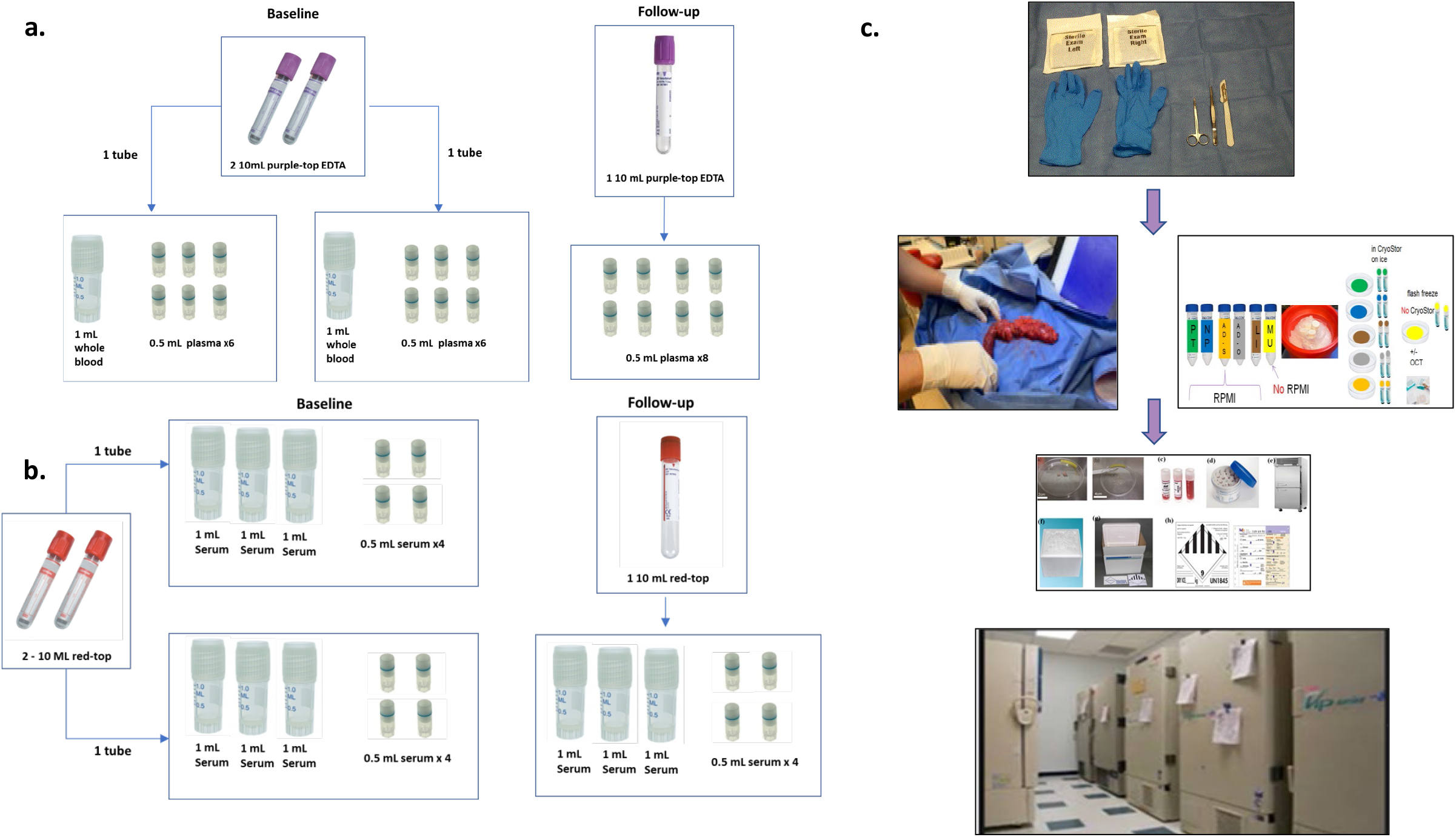
Blood and Tissue Collection and Processing Workflow. a) Purple-topped EDTA tubes are processed and stored at Baseline and Follow-up time-points into whole blood (Baseline) and plasma (Baseline and Follow-up). b) Red-topped tubes are processed and stored at Baseline and Follow-up timepoints into serum. c) During the surgical procedure, a debridement kit containing sterile supplies (gloves, gauze, scissors, forceps, scalpel, and drape) is open in preparation for receiving tissue samples. Collected tissue samples are placed in labelled conical tubes on ice with 20mL RPMI 1640/2% penicillin-streptomycin (Pancreatic Tumor (PT), Normal Pancreas (NP), Adipose – omental (AD-O), Adipose – subcutaneous (AD-S), and Liver (LI), or in an empty conical tube Muscle (MU). Samples are then transferred to labelled petri dishes for mincing. Non-muscle tissues are placed in cryovials with CryoStor CS10 to slow freeze in a Mr. Frosty overnight prior to long-term storage in liquid nitrogen. Muscle tissues are either placed in cryovials with no media and snap frozen prior to long-term storage in a −80°C freezer (Step A) or frozen in isopentane, embedded in OCT, and stored in a cassette in a −80°C freezer until shipment and future analysis (Step B).

##### Surgically-resected tissue

At the time of surgical resection, sites collect and process pancreatic tumor (PT), normal pancreas (NP), and muscle (MU) from the upper right quadrant of the rectus abdominus, adipose-subcutaneous above rectus muscle (AD-S), adipose-omental/intraperitoneal (AD-O), and sites of metastasis such as liver (LI). As shown in **Figure 2c**, a debridement kit containing sterile supplies (gloves, drape, forceps, scissors, scalpel, and gauze) is provided for use in the pathology gross room. Sites organize supplies with color-coded cryodots corresponding to each tissue type, including pre-labeled 50 mL conical tubes containing 20mL RPMI 1640/2% penicillin-streptomycin (p/s), petri dishes, and 2 mL cryovials (Argos Polarsafe Cryogenic Storage Vials with External Cap), filled with 1 mL of CryoStor CS10 freezing solution (BioLife Solutions, Inc) for all tissue types except for MU. (MU is not placed into RPMI or CryoStor.) For each tissue type, the time of tissue removal, time received in pathology suite (for PT & NP only), time placed in media, and time of freezing is recorded. After tumor resection, the time until tissue immersion into preservative is typically under 30 minutes to minimize ischemia and degradation.

When the resected pancreatic tumor specimen is removed, the surgeon immediately obtains an en face section of the pancreatic transection margin for frozen-section and the specimen is transported on wet ice for gross analysis by a pathologist and/or pathology assistant. The local pathology team determines whether ample pancreatic tumor tissue is available for diagnosis and cancer staging and whether a portion of the specimen can be harvested for banking without disrupting the accuracy of the pathology reporting for patient care. The priority for banking is the central area of the tumor followed by the tumor margin, and a minimum of a 5.0 mm^3^ tumor fragment from the epicenter is requested. Upon obtaining a negative margin on the pancreatic edge, we aim to obtain a “*normal*” pancreatic biopsy at the reconstructive end. The site priority is (in decreasing order): distant pancreas, grossly uninvolved pancreas, perilesional uninvolved pancreas (normal tissue adjacent to the tumor, or surrounding stroma). Non-MU tissue samples are placed in the corresponding conical tube on ice and transferred to a designated institutional laboratory/processing facility where sterile forceps are used to transfer each tissue type to the corresponding pre-labeled petri dish along with 3 mL of RPMI with 2% p/s from each sample’s conical tube. Non-muscle samples (PT, NP, AD-S, AD-O and any metastatic specimens) are minced into 2-3mm^3^ fragments using sterile forceps and scalpels and 2-6 fragments are transferred to pre-labeled Cryovials preloaded with 1 mL CryoStor CS10 freezing solution. Cryovials are immediately stored at 4°C for 30 min to allow CryoStor solution to penetrate the tissue and then placed into a Mr. Frosty Freezing Container (ThermoFisher Scientific) (4°C) and stored at −80°C overnight. Cryovials are transferred to a liquid nitrogen (LN2) storage unit, vapor phase or shipped to the coordinating center the next day per SOPs **(Figure 2c)**. Tissue is stored for future applications and translation research efforts including generation of cell lines, organoids, or xenografts^79-83^.

Two main steps are involved in processing MU tissue: flash freezing for future biochemical analysis (step A) and embedding in optimal cutting temperature (OCT) compound for future morphological assessment (step B). Upon incision through the skin and dissection through subcutaneous fat, a 2.0 × 1.0 cm muscle biopsy specimen is obtained from the upper right quadrant of the rectus abdominis and sharply divided into four fragments, avoiding cautery burns. Three of the 4 fragments are placed into three pre-labeled cryovials and put in a LN2-containing dewar (or dry ice/ethanol slurry if liquid nitrogen is not available) and stored in a −80°C freezer for step A. The fourth fragment (used for step B) is wrapped in gauze pre-moistened with ice-cold PBS, placed in a pre-labeled conical tube on *wet* ice, embedded in OCT, frozen in liquid nitrogen-cooled isopentane, cooled in liquid nitrogen, and stored at −80°C until shipped on dry ice to the UFG site for storage and future analysis by our team’s muscle physiologists (AJ, SJ).

##### Endoscopic fine needle aspirate and core biopsies

In September of 2019, we began collection of cystic fluid and tissue from patients undergoing endoscopic ultrasound-guided fine needle aspirate (FNA) and fine needle aspirate biopsies (FNAB) from cystic and solid pancreatic neoplasms, respectively. Initial passes are designated for diagnostic purposes, and up to three additional passes are collected for research if deemed safe by the endoscopists. Residual cystic fluid over 2 mL is aliquoted into 4 × 1 ml digital barcode-labeled cryovials with 0.5 mL of sample per cryovial and stored at –80°C. FNAB smears and cell blocks are prepared according to institutional cytology laboratory standards. The FNAB needle is rinsed in 5–10 mL of balanced salt solution or other medium. This sample is then centrifuged, and the pellet used to prepare a cell block. The residual supernatant is saved and stored at −80°C until shipped to MCC.

##### Repository of CT images

Consistent with the missions of NCI’s Quantitative Imaging Network^84^ and Cancer Imaging Archive (TCIA)^85^, we use best practices for acquisition, de-identification, curation, and secure transmission/sharing of pancreas-specific radiologic images with focus on CT images. TCIA has SOPs for Digital Imaging and Communications in Medicine (DICOM) images and metadata that are being followed^85^. Participating sites provide CT scans at each timepoint from consented participants to Moffitt’s Quantitative Imaging (QI) Team via CD or electronically to a secure ShareFile or Powershare portal. Instructions for DICOM header re-labeling and transmission are followed and tracking surveys are completed in DatStat. Upon receipt, the QI Team uploads the corresponding imaging report and CT scans into MCC’s Healthmyne Research Infrastructure and logs scan details into an Excel database. Additional metrics are abstracted and entered into a standardized template for radiologic reporting of PDAC^86^. Donated images are also being used to perform body composition analyses.

##### Development of an Integrated and Centralized Virtual Data Repository

A central database linking individual-level de-identified data to biospecimens and images across various internal and external source systems has been created and maintained by the investigative team at MCC and is known as the Florida Pancreas Collaborative Data Repository (FPCDR). Outside of the demilitarized zone (DMZ), the virtual repository integrates electronic survey data ascertained through DatStat with paper survey data provided through MCC’s Participant Research, Interventions, and Measurement (PRISM) Core, data from the STRAIN^72^ instrument housed on a server at the University of California, and cancer registry data from the FCDS **(Supplementary Figure 6)**. The study web-site and the ShareFile application used to transfer de-identified images and reports is also housed outside of the DMZ until joined to participant information and stored on the MCC network as part of the image repository, DatStat database, or other source systems. For example, demographic data is transferred using HL7 to MCC’s Clinical Trails Management System (Oncore), and biospecimen-level annotation is transferred into LabVantage and includes variables such as the date of collection, tissue of origin, primary site of disease, histological diagnosis, storage format (i.e. LN2, OCT, −80°C), the number of cryovials vials of each sample in each storage format (box, row, and vial number) and their location. Reporting and querying functions are currently being developed to help with generating summary reports and ad hoc queries. In addition to the security in place through the DatStat platform itself (which include including Health Insurance Portability and Accountability Act (HIPAA) compliance, 21 CFR Part 11 Compliance, SOC 2 Type 2 Certification, and Privacy Shield certification), numerous safeguards have been incorporated into the centralized repository to maintain patient confidentiality and ensure compliance with HIPAA guidelines. Data also undergo quality and post-load checks periodically to identify incorrect or missing data.

##### Descriptive Statistics

Frequencies and percentages were generated for categorical variables and means and standard deviations (SD) were calculated for continuous variables. Distributions of covariates were compared across racial/ethnic groups using chi-squared tests, fisher’s exact tests, t-tests, and generalized linear models. Pvalues <0.05 were considered statistically significant. Categorization of the cachexia continuum and depression were performed using established methodology^87,88^. All analyses were performed using SAS version 9.4 (SAS Institute, Cary, North Carolina).

## Results

### Enrollment

Recruitment was initiated at MCC in March 2019. As of August 2020, 13 of our 15 FPC sites were actively recruiting with 2 sites delayed due to finalizing the site agreement (n=1) and institutional obstacles including turnover of key study staff (n=1). A total of 350 individuals (264 NHW, 32 AA, 53 H/L and 1 unknown race/ethnicity) were identified and assessed for eligibility to participate. Of 323 individuals deemed to be eligible (243 NHW, 30 AA, and 50 H/L), 305 enrolled (228 NHW, 30 AA, and 47 H/L), with participation rates of 94%, 100%, and 94%, respectively. Nearly 41% (n=124) of enrolled participants were recruited at sites other than the three major academic cancer centers in the state (MCC, UFG, and UOM) **(Supplementary Figure 7)**. A detailed flowchart regarding recruitment outcomes including reasons for exclusion and for declining enrollment is shown in **Figure 3**.

**Figure 3.**
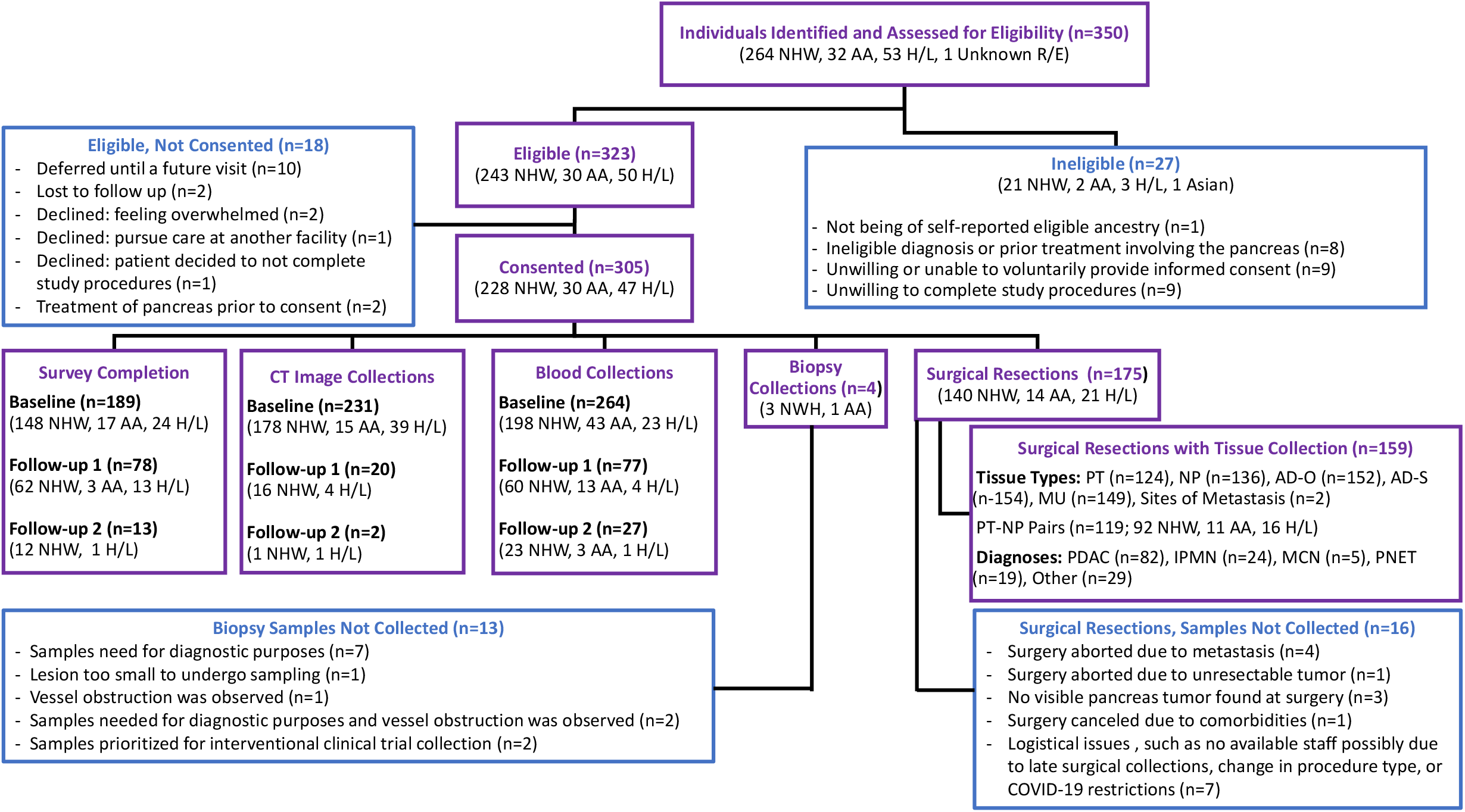
Summary of Recruitment, Survey Data, Image, and Biospecimen Collection Efforts to Date, by Race/Ethnicity. The flow diagram depicts the number of individuals eligible and ineligible for the study, as well as number of consented participants who have donated biospecimens, computed tomography (CT) images, and completed surveys. Abbreviations: Non-Hispanic White (NHW), African American (AA), Hispanic/Latinx (H/L), Pancreatic Tumor (PT), Normal Pancreas (NP), Adipose – omental (AD-O), Adipose – subcutaneous (AD-S), Muscle (MU), Pancreatic Ductal Adenocarcinoma (PDAC), Intraductal Papillary Mucinous Neoplasm (IPMN), Mucinous Cystic Neoplasm (MCN), Pancreatic Neuroendocrine Tumor (PNET).

### Study population characteristics

Select characteristics of the study population are summarized in **Table 3**. The average age at diagnosis in our study cohort is 68 years, with AA and H/L having significantly younger ages at diagnosis than NHW (64 and 63 versus 70 years, respectively, P=0.0001). Most study participants (n=161, 53%) are female, with the highest proportion of females observed among H/L. Preliminary data on education and income level and health insurance status did not identify significant differences between racial/ethnic groups. The most common presenting symptoms included weight loss of >5% over the past 6 months (n=115, 40.9%), abdominal pain (n=100, 49.8%), fatigue (n=98, 34.7%) and jaundice (n=66, 23.3%). Mean BMI was highest among AA (28 kg/m^2^) followed by NHW (27 kg/m^2^) and H/L (25 kg/m^2^). Almost one-third of the study population (n=90) reported a personal history of diabetes, with no statistically significant differences between racial/ethnic groups. Most participants have a confirmed diagnosis of PDAC (n=183, 61.4%), followed by IPMNs and PNET, each representing 11.7% (n=35) of cases. Seven of the AA cases with confirmed histology (25%) had PNETs, which is significantly higher than the proportion of PNETs identified in NHW (11.1%) and H/L (6.5%). In a sensitivity analysis comparing all PDAC and PNET cases, it was observed that PNET cases in the cohort were: more likely to be diagnosed at a younger age (63 vs. 69 years, P=0.0022); have higher incomes (P=0.0173); and less likely to have presented with jaundice and >5% weight loss (P=0.0054 and P=0.0267, respectively) (data not shown). Of the 183 PDAC cases, staging data is available for 99 (60 (32.8%) are stage I/II and 39 (21.3%) are stage III/IV). Of the 22 PNET cases for which staging data is available, 54.3% are stage I/II and 2.9% are stage III/IV (P=0.0001). Overall, based primarily on patient reported outcomes in the health screen and baseline questionnaire, the prevalence of cachexia, depression, former tobacco use, and current tobacco use upon study enrollment were estimated to be 32.9%, 35.1%, 43.6%, and 12.1% respectively.

**Table 3.** Baseline demographic and clinical characteristics of Florida Pancreas Collaborative study participants, by race/ethnicity.

| Variable | All Participants<br>(N=305) | African American (N=30) | Hispanic/Latinx<br>(N=47) | Non-Hispanic White<br>(N=228) | P-Value |
| --- | --- | --- | --- | --- | --- |
| Age (years), mean (+/- SD) | 68 (10.6) | 64 (12.1) | 63 (12.8) | 70 (9.4) | 0.0001 |
| Gender, n (%) |  |  |  |  |  |
| Female | 161 (52.8) | 17 (56.7) | 30 (63.8) | 114 (50%) | 0.2028 |
| Male | 144 (47.2) | 13 (43.3) | 17 (36.2) | 114 (50%) |  |
| Education level<sup>†</sup>, n(%) |  |  |  |  |  |
| High school or GED | 46 (31.1) | 6 (54.5) | 6 (42.9) | 34 (27.6) | 0.1599 |
| College | 65 (43.9) | 4 (36.4) | 7 (50.0) | 54 (43.9) |  |
| Post graduate | 37 (25.0) | 1 (9.1) | 1 (7.1) | 35 (28.5) |  |
| Data not yet available<sup>§</sup> | 157 | 19 | 33 | 105 |  |
| Income Level<sup>†</sup>, n (%) |  |  |  |  |  |
| Below \$40k | 38 (26.0) | 6 (54.5) | 4 (30.8) | 28 (22.9) | 0.2049 |
| \$40k-100k | 42 (28.8) | 2 (18.2) | 3 (23.1) | 37 (30.3) | |
| 100k and above | 34 (23.3) | 1 (9.1) | 1 (7.7) | 32 (26.3) |  |
| Information not provided by Participant | 32 (21.9) | 2 (18.2) | 5 (38.5) | 25 (20.5) |  |
| Data not yet available<sup>§</sup> | 159 | 19 | 34 | 106 |  |
| Health Insurance<sup>†</sup>, n (%) |  |  |  |  |  |
| Insured | 143 (97.9) | 10 (90.9) | 13 (100.0) | 120 (98.4) | 0.2301 |
| Uninsured | 3 (2.1) | 1 (9.1) | 0 (0.0) | 2 (1.6) |  |
| Data not yet available<sup>§</sup> | 159 | 19 | 34 | 106 |  |
| Marital Status<sup>†</sup>, n (%) |  |  |  |  |  |
| Not married | 38 (26.0) | 5 (45.5) | 2 (15.4) | 31 (25.4) | 0.0376 |
| Married | 107 (73.2) | 5 (45.5) | 11 (84.6) | 91 (74.6) |  |
| Information not provided by Participant | 1 (0.8) | 1 (9.0) | 0 (0.0) | 0 (0.0) |  |
| Data not yet available | 159 | 19 | 34 | 106 |  |
| Family History of Pancreatic Cancer<sup>†</sup>, n (%) |  |  |  |  |  |
| No | 87 (67.4) | 8 (72.7) | 6 (66.7) | 73 (67.0) | 0.8005 |
| Yes | 16 (12.4) | 0 (0.0) | 1 (11.1) | 15 (13.8) |  |
| Participant does not know | 26 (20.2) | 3 (27.3) | 2 (22.2) | 21 (19.2) |  |
| Data not yet available<sup>§</sup> | 176 | 19 | 38 | 119 |  |
| Distress<sup>†</sup>, n (%) |  |  |  |  |  |
| No | 36 (12.4) | 3 (11.1) | 7 (14.9) | 26 (12.0) | 0.8309 |
| Yes | 255 (87.6) | 24 (88.9) | 40 (85.1) | 191 (88.0) |  |
| Data not yet available<sup>§</sup> | 14 | 3 | 0 | 11 |  |
| Depression<sup>†</sup>, n (%) |  |  |  |  |  |
| No | 189 (64.9) | 20 (74.1) | 33 (70.2) | 136 (62.7) | 0.3579 |
| Mild depression | 43 (14.8) | 4 (14.8) | 3 (6.4) | 36 (16.6) |  |
| Moderate depression | 40 (13.8) | 1 (3.7) | 7 (14.9) | 32 (14.7) |  |
| Severe depression | 19 (6.5) | 2 (7.4) | 4 (6.4) | 13 (6.0) |  |
| Data not yet available<sup>§</sup> | 14 | 3 | 0 | 11 |  |
| Smoking status<sup>†‡</sup>, n (%) |  |  |  |  |  |
| No | 129 (44.3) | 14 (50) | 32 (68.1) | 83 (38.4) | 0.0055 |
| Former smoker | 127 (43.6) | 10 (35.7) | 12 (25.5) | 105 (48.6) |  |
| Current smoker | 35 (12.1) | 4 (14.3) | 3 (6.4) | 28 (13.0) |  |
| Data not yet available<sup>§</sup> | 14 | 2 | 0 | 12 |  |
| Marijuana status<sup>†</sup>, n (%) |  |  |  |  |  |
| No | 97 (71.9) | 5 (50) | 10 (90.9) | 82 (71.9) | 0.1662 |
| Former user | 25 (18.5) | 3 (30) | 0 (0.0) | 22 (19.3) |  |
| Current user | 13 (9.6) | 2 (20) | 1 (9.1) | 10 (8.8) |  |
| Data not yet available<sup>§</sup> | 170 | 20 | 36 | 114 |  |
| Abdominal Pain<sup>†</sup>, n (%) |  |  |  |  |  |
| No | 78 (38.8) | 12 (46.2) | 11 (31.4) | 55 (39.3) | 0.0948 |
| Yes | 100 (49.8) | 13 (50.0) | 23 (65.7) | 64 (45.7) |  |
| Information unavailable in EMR | 23 (11.4) | 1 (3.8) | 1 (2.9) | 21 (15.0) |  |
| Data not yet available<sup>§</sup> | 104 | 4 | 12 | 88 |  |
| Fatigue<sup>†</sup>, n (%) |  |  |  |  |  |
| No | 148 (52.5) | 15 (53.6) | 20 (45.5) | 113 (53.8) | 0.0975 |
| Yes | 98 (34.7) | 12 (42.8) | 21 (47.7) | 65 (31.0) |  |
| Information unavailable in EMR | 36 (12.8) | 1 (3.6) | 3 (6.8) | 32 (15.2) |  |
| Data not yet available<sup>§</sup> | 23 | 2 | 3 | 18 |  |
| GI Bleeding<sup>†</sup>, n (%) |  |  |  |  |  |
| No | 217 (77.0) | 23 (82.1) | 39 (88.6) | 155 (73.8) | 0.0346 |
| Yes | 7 (2.5) | 2 (7.1) | 1 (2.3) | 4 (1.9) |  |
| Information unavailable in EMR | 58 (20.5) | 3 (10.8) | 4 (9.1) | 51 (24.3) |  |
| Data not yet available<sup>§</sup> | 23 | 2 | 3 | 18 |  |
| Jaundice<sup>†</sup>, n (%) |  |  |  |  |  |
| No | 178 (62.9) | 22 (78.6) | 25 (55.6) | 131 (62.4) | 0.0129 |
| Yes | 66 (23.3) | 4 (14.3) | 18 (40.0) | 44 (21.0) |  |
| Information unavailable in EMR | 39 (13.8) | 2 (7.1) | 2 (4.4) | 35 (16.6) |  |
| Data not yet available<sup>§</sup> | 22 | 2 | 2 | 18 |  |
| Weight Loss More Than 5%<sup>†</sup>, n (%) |  |  |  |  |  |
| No | 133 (47.4) | 22 (78.6) | 25 (55.6) | 131 (62.4) | 0.0972 |
| Yes | 115 (40.9) | 4 (14.3) | 18 (40.0) | 44 (21.0) |  |
| Information unavailable in EMR | 33 (11.7) | 2 (7.1) | 2 (4.4) | 35 (16.6) |  |
| Data not yet available<sup>§</sup> | 24 | 2 | 2 | 18 |  |
| Charlsons Comorbidity Index, n (%) |  |  |  |  |  |
| 0 | 164 (57.7) | 16 (57.1) | 27 (60.0) | 121 (57.3) | 0.6516 |
| <=2 | 101 (35.6) | 11 (39.3) | 13 (28.9) | 77 (36.5) |  |
| >=3 | 19 (6.7) | 1 (3.6) | 5 (11.1) | 13 (6.2) |  |
| Data not yet available<sup>§</sup> | 21 | 2 | 2 | 17 |  |
| Personal History of Diabetes<sup>††</sup>, n (%) |  |  |  |  |  |
| No | 195 (68.4) | 21 (75) | 27 (60) | 147 (69.3) | 0.3464 |
| Yes | 90 (31.6) | 7 (25) | 18 (40) | 65 (30.7) |  |
| Data not yet available<sup>§</sup> | 20 | 2 | 2 | 16 |  |
| Personal History of Pancreatitis<sup>††</sup>, n (%) |  |  |  |  |  |
| No | 180 (79.6) | 23 (85.2) | 32 (86.5) | 125 (77.2) | 0.3745 |
| Yes | 46 (20.4) | 4 (14.8) | 5 (13.5) | 37 (22.8) |  |
| Data not yet available<sup>§</sup> | 79 | 3 | 10 | 66 |  |
| Cachexia<sup>††</sup>, n (%) |  |  |  |  |  |
| refractory cachexia | 1 (3.8) | 0 (0.0) | 4 (8.9) | 6 (3.1) | 0.3098 |
| cachexia | 76 (29.1) | 4 (18.2) | 16 (35.6) | 56 (28.9) |  |
| pre-cachexia | 26 (10.0) | 4 (18.2) | 2 (4.4) | 20 (10.3) |  |
| non cachectic | 149 (57.1) | 14 (63.6) | 23 (51.1) | 112 (57.7) |  |
| missing | 44 | 8 | 2 | 34 |  |
| Body Mass Index (kg/m2)<sup>†</sup><br>n, mean (SD) | 281, 27 (5.5) | 26, 28 (5.2) | 44, 25 (5.0) | 211, 27 (5.5) | 0.0348 |
| Waist Circumference,<sup>†</sup><br>n, mean (SD) | 231, 40 (12.7) | 17, 39 (6.2) | 37, 38 (13.5) | 177, 40 (13.0) | 0.6594 |
| Histology<sup>†</sup>, n (%) |  |  |  |  |  |
| Pancreatic ductal adenocarcinoma (PDAC) | 183 (61.4) | 16 (57.2) | 36 (78.3) | 131 (58.5) | 0.0200 |
| Pancreatic neuroendocrine tumor (PNET) | 35 (11.7) | 7 (25.0) | 3 (6.5) | 25 (11.1) |  |
| Intraductal papillary mucinous neoplasm (IPMN) | 35 (11.7) | 1 (3.6) | 2 (4.3) | 32 (14.3) |  |
| Mucinous cystic neoplasm (MCN) | 6 (2.0) | 2 (7.1) | 0 (0.0) | 4 (1.8) |  |
| Other<sup>§</sup> | 39 (13.1) | 2 (7.1) | 5 (10.9) | 32 (14.3) |  |
| Data not yet available<sup>§</sup> | 7 | 2 | 1 | 4 |  |
| Surgical Resection Attempted<sup>†</sup>, n (%) |  |  |  |  |  |
| No | 146 (47.9) | 17 (56.7) | 26 (55.3) | 103 (45.2) | 0.2673 |
| Yes | 159 (52.1) | 13 (43.3) | 21 (44.7) | 125 (54.8) |  |
| Location of Tumor<sup>†</sup>, n (%) |  |  |  |  |  |
| Body | 16 (14.0) | 2 (33.3) | 1 (4.8) | 13 (13.3) | 0.3351 |
| Diffuse | 17 (14.9) | 2 (33.3) | 3 (14.3) | 12 (12.4) |  |
| Head | 65 (57.0) | 1 (16.7) | 13 (61.8) | 51 (52.6) |  |
| Tail | 13 (11.4) | 0 (0.0) | 1 (4.8) | 12 (12.4) |  |
| Other | 13 (11.4) | 1 (16.7) | 3 (14.3) | 9 (9.3) |  |
| Data not yet available<sup>§</sup> | 181 | 24 | 26 | 131 |  |
| Stage<sup>†</sup>, n (%) |  |  |  |  |  |
| Stage 0 | 22 (14.1) | 1 (20.0) | 1 (4.5) | 20 (16.3) | 0.6547 |
| Stage I/II | 87 (55.8) | 3 (60.0) | 14 (63.7) | 70 (56.9) |  |
| Stage III/IV | 41 (26.3) | 1 (20.0) | 7 (31.8) | 33 (26.8) |  |
| Data not yet available<sup>§</sup> | 155 | 25 | 25 | 105 |  |
| Grade Exocrine Pancreatic Tumors<sup>†††</sup>, n (%) |  |  |  |  |  |
| Well differentiated | 7 (8.0) | 0 (0.0) | 3 (15.8) | 4 (6.1) | 0.1719 |
| Moderately differentiated | 29 (32.9) | 0 (0.0) | 9 (47.4) | 20 (30.3) |  |
| Poorly differentiated | 21 (23.9) | 1 (33.3) | 4 (21.0) | 16 (24.2) |  |
| Grade undetermined | 31 (35.2) | 2 (66.7) | 3 (15.8) | 26 (39.4) |  |
| Data not yet available<sup>‡</sup> | 136 | 16 | 19 | 101 |  |
| Grade IPMN<sup>¶</sup>, n (%) |  |  |  |  |  |
| Low grade | 12 (34.3) | 0 (0.0) | 1 (50.0) | 11 (34.4) | 1.0000 |
| Borderline | 2 (5.7) | 0 (0.0) | 0 (0.0) | 2 (6.3) |  |
| Carcinoma-in-situ | 5 (14.3) | 0 (0.0) | 0 (0.0) | 5 (15.6) |  |
| Invasive carcinoma | 1 (2.9) | 0 (0.0) | 0 (0.0) | 1 (3.1) |  |
| Unknown grade | 15 (42.9) | 1 (100.0) | 1 (50.0) | 13 (40.6) |  |
| Positive Lymph Nodes<sup>§</sup>, n (%) |  |  |  |  |  |
| No | 110 (94.8) | 5 (100.0) | 17 (89.5) | 88 (95.7) | 0.6800 |
| Yes | 6 (5.2) | 0 (0.0) | 2 (10.5) | 4 (4.3) |  |
| Data not yet available<sup>‡</sup> | 189 | 25 | 28 | 136 |  |

<sup>†</sup>Data available from partially-or fully-completed baseline questionnaires at time of analysis.
<sup>‡</sup>Data not yet available. Additional data will be included in future analyses when entered into the DatStat system.
<sup>‡</sup>Data available from the health screen questionnaire at time of analysis.
<sup>¶</sup>Data available from case report forms (CRFs) at time of analysis.
<sup>§</sup>The 'Other' category includes benign and malignant tumors of pancreatic, liver and bile duct, renal, adrenal gland, lymph node, and unclassified origin.
<sup>\*</sup>The grade for exocrine tumors has been restricted to PDAC, IPMN, and MCN (n=224).

### Exited Participants

Forty-seven participants have been exited or withdrawn from the study thus far. Reasons include death (n=28), screen failure/found to be ineligible (n=8), withdrawal by the participant (n=6), treated at another institution (n=3), admission to hospice (n=1), and withdrawal by the physician (n=1). Of those who died, most (n=25, 89.2%) had PDAC, 2 had chronic cholecystitis, and 1 has an unknown diagnosis. A sensitivity analysis comparing characteristics of exited participants and those not exited did not reveal any significant differences in sociodemographic factors; most exited participants had stage III/IV PDAC and have died (**Supplementary Table 2**).

### Survey completion

Over an 18-month period, 189 baseline surveys have been completed (62% completion rate). Baseline survey completion was higher for participants with PNET (77.1%) compared to PDAC (56.8%). Of the 189 participants who have completed the baseline survey, 164 have reached follow-up 1 (6-month visit) and 76 completed that survey (46% completion rate). To increase completion rates, participants receive automated email and/or phone call reminders from site coordinators to complete the questionnaires.

### Computed tomography (CT) scan acquisition

Baseline CT images from 231 participants (178 NHW, 15 AA, 38 H/L) have been uploaded to our central imaging repository **(Figure 3)**. Most scans are ‘CT Abdomen Pelvis’ (n=73, 32%). ‘CT Abdomen’ and ‘CT Thorax Abdomen Pelvis’ account for 26% of these scans (n=60), followed by ‘Pancreas Protocol CT’ scans (n=30, 13%). An additional 26% of CT scans are of different types such as PET/CT. CT scans from follow-up time points 1 and 2 have been received for 20 and 2 participants, respectively.

### Biospecimen collection

As summarized in **Figure 3**, blood samples have been obtained at baseline for 264 participants (198 NHW, 23 AA, 43 H/L) and at follow-up timepoints 1 and 2 for 77 and 27 participants, respectively. Tissue samples have been collected from 159 of 175 surgical participants, with 119 matched PT and NP pairs, 152 AD-O, and 149 MU samples collected. Most pancreatic tumor samples (n=114, 91.9%) were collected prior to any treatment, while ten samples were collected post-treatment. Of the 159 participants with available tissue samples, most had a diagnosis of PDAC (52%, n=82) followed by IPMNs (15%, n=24) and PNETs (12%, n=19).

## Discussion

In this report we describe the establishment of the first state-wide biobank dedicated to minimizing disparities and personalizing care for individuals affected by PaCa. Throughout this multiple stakeholder-led initiative, we have proactively developed and implemented robust SOPs to collect, process, and store blood and tissues uniformly to ensure quality specimens for downstream analyses. Moreover, we developed standardized methods for the collection of demographic, socioeconomic, clinical, laboratory, and epidemiologic data with which to annotate the biospecimens. By integrating data with biomarkers derived from biospecimens and medical images, we hope to investigate biological processes that may underlie disparities and poor outcomes and develop targeted and personalized interventions that may improve outcomes for individuals. In line with prior studies of PaCa disparities^4,6-11,38,89^, the AA cases enrolled in our cohort to date were diagnosed at younger ages (mean=64 years) and have a higher BMI than NHW. In contrast to retrospective reviews of cancer registry data from other large stated including Texas^8^ and California^10^, we are observing a higher proportion of females affected by PaCa in our cohort, especially among AA and H/L.

It is well documented that recruitment and enrollment of underserved populations into cancer biobanks and clinical trials is challenging due to multiple factors: lack of trust; concerns surrounding protection of privacy; language barriers; aversion to additional blood draws; transportation issues; and institutional barriers including organizational challenges and lack of resources to conduct research^90-96^. The FPC has worked diligently to address these barriers through the incorporation of known facilitators, including use of Spanish-translated written materials, combining study visits and blood draws with standard of care appointments, and enlisting the promotion of the study by engaged providers trusted by potential participants^95,97^. To address institutional barriers, the FPC coordinating center has worked closely with academic and community hospitals to meet with pathologists, laboratory technicians, regulatory specialists, and business offices to obtain buy-in for the study, ensure a study coordinator is available for study visits and data entry, and that all study-related supplies (including iPads) are provided. This institutional support has been essential to ensuring the study is accessible to populations seen at lower-volume hospitals and/or those hospitals without research infrastructure in place.

The 30 AA and 47 H/L individuals recruited to date account for 9.8% and 15.4% of total participants recruited, respectively, representing percentages higher than those reported in existing pancreas biobanks^46,47^ and in existing molecular studies of PaCa and other pancreatic tumors from TCGA and other initiatives^98-102^. A summary comparing the unique FPC resource with the PDAC TCGA cohort (referred to as ‘PAAD’)^102^ underscores how the FPC is filling important gaps in PaCa disparities research by: including a greater representation of minority groups and collecting numerous untreated and treated tissue types in addition to pancreas tumor tissue along with CT scans, blood, and a comprehensive set of clinical, epidemiologic, laboratory, and quality of life data elements (**Supplementary Figure 8**). Importantly, compared to other biospecimen donation studies^90,103,104^, the willingness of eligible AA and H/L patients to participate has been remarkably high at 100% and 94%, respectively. Thus, despite what may appear to be relatively small sample sizes to date, the FPC has been experiencing success enrolling underserved populations at sites selected based on state data^49^. Therefore, with continued FPC activities, we expect the numbers of AA and H/L cases to increase further, particularly with the activation of our two remaining sites which together see a high volume of both AA and H/L patients. Furthermore, we expect to accelerate enrollment as remote recruitment efforts are increasingly adopted by sites in response to the COVID-19 pandemic. The FPC recognizes challenges associated with accessing underserved populations throughout the state who may be pursuing care at lower-volume hospitals and facilities that do not facilitate surgery or chemotherapy when appropriate. We are actively pursuing opportunities to engage and educate additional providers that may be able to bridge this gap and ensure appropriate treatment is provided to incident cases.

In terms of immediate plans, we will be working to advance cancer cachexia research using data elements collected via the study questionnaire and health screen, laboratory values from the EMR, and correlates of cancer cachexia characterized by quantitative imaging features derived from CT scans (such as metrics of visceral adiposity, skeletal muscle index, and psoas muscle index) and molecular markers (such as cytokines and adipokines) measured using serum samples. We will also be performing genotyping for ancestry informative markers using DNA from blood collected via this study to validate self-reported ancestry. Finally, based on the promise of pre-clinical models in translational efforts^79-83^, we also plan to leverage the pancreatic tissue collected and preserved through this effort for applications such as generation of patient-derived organoids^105^. Thus, the biospecimens collected by the FPC will become an enduring resource for the biomedical and disparities research community.

Investigators in and outside the FPC may request to collaborate and utilize these resources to evaluate new research hypotheses after a series of analyses are conducted and published by the FPC. A written proposal would be submitted and reviewed by the FPC Biobank Utilization Committee with decisions made based upon peer-review of scientific merit, specimen availability, experience of the requesting investigator(s), and resource adequacy to conduct proposed methods. Samples and/or data are released upon committee approval once the requestors secure regulatory approvals, conflicts of interest disclosures are reviewed, and data use and material transfer agreements are established. Intellectual property issues would be agreed upon in advance with results from the new research findings incorporated into the biobank. In this manner, the FPC data repository continues to evolve, generating new correlations and opportunities to evaluate hypotheses.

In summary, multidisciplinary, multi-institutional collaborations in partnership with community stakeholders will be key to successfully addressing PaCa disparities. Importantly, Boyd and colleagues^44^ suggest that closing the gap in racial health outcomes will also necessitate ‘identifying, confronting, and abolishing racism as an American tradition and root of inequity.’ Taken together, it is critical that community and institutional leaders and other stakeholders support and enable these important cancer disparities initiatives. It is our intent that the infrastructure-building described here can serve as a model for other teams who wish to develop similar resources applicable to their disease sites.

## Data Availability

Data for this manuscript is not publicly available. It is protected in a secure database by the lead investigator.

## Acknowledgements

We thank our participants and Community Advisory Board Members Lorriel S. Blaise and the late Suzy Swenson for their support and dedication to this project. We also thank the following FPC team members for their collaboration and contributions to generating original data and/or data abstraction and entry: Lidia Sanchez, Jennifer Mull, Yohan Zuniga, Ma Therea De Leon, Lisa Rivero, Rebecca Delph, Brittney Johnson, Yvonne Zuniga, Kim Lacy, Jon-Michael Eckert, Khattiya Chharath, Beth Montera, Thanh Tran, Alexandra Tassielli, Tri Huynh, and Thinzar Zaw. The research in this publication was supported in part by the James and Esther King Biomedical Research Program, Florida Department of Health (Grant #8JK02; awarded to JPB and JGT), and the Tissue Core, Quantitative Imaging Core, the Participant Research, Interventions, and Measurement (PRISM) Core, the Biostatistics and Bioinformatics Shared Resource, and the Pharmacokinetics and Pharmacodynamics Core at the H. Lee Moffitt Cancer Center & Research Institute, an NCI designated Comprehensive Cancer Center (P30-CA076292). The content is solely the responsibility of the authors and does not necessarily represent the official views of the sponsors or the H. Lee Moffitt Cancer Center & Research Institute. The authors also wish to thank Warren Gloria, Karen Coley, Rithi Somesh Shivaram, Dennis Hall, and Vyacheslav Petrovsky for their assistance with pathological review of tissues in the gross room.

---

**Supplementary Figure 1. Workflow of Main Study-related Tasks Accomplished with the Assistance of our Online Data Management/Engagement Platform**.

Workflow includes screening patients to recruit, registering their demographic information and language preference (English or Spanish), automatic assignment of a unique identification number (ID) which is distinct from their medical record numbers and has a three-letter site prefix followed by four numbers (i.e. MCC-1234), recording approach outcomes, assessment of eligibility, obtaining informed consent electronically or documenting paper consent, verification of contact information for the participant and at least one individual who can be contacted if the participant cannot be reached, tracking of the collection, processing, transfer, and storage of biospecimens and images, sending automated email notifications related to study tasks, administration of questionnaires/surveys, and report generation. Abbreviations: EMR=electronic medical record; PHI=protected health information; E=electronic; CRF=case report form.

**Supplementary Figure 2. Workflow and Modules Represented in the Electronic-Consent (e-consent) Process**.

The E-Consent Process has an array of components that allow for more efficient recruitment. This process is deconstructed into multiple modules: a) Add Participant Form, b) Eligibility Form, c) Approach Attempts, d) Name and Contact Information Form, and e) an informed consent form (ICF). Upon first screening a patient, the site coordinator completes the Add Participant form thereby collecting non-protected health information (PHI). Upon completion of that module, the coordinator is automatically directed to the Eligibility Form which then assesses the patient’s eligibility through a series of questions. If eligible, the coordinator then chooses the next module depending on the outcome of the approach. If the patient agrees to consent, a Contact Information Module automatically populates and directly feeds all PHI into the E-Consent. Within the E-Consent, the patient electronically signs all portions of the document, including permission to contact a relative of their choosing if they cannot be reached during their study involvement. On the other hand, if the patient defers or declines, the coordinator is prompted to the Approach Attempts Module and Exit Module, respectively, where specific reasons for can be documented and pulled for future encounters.

**Supplementary Figure 3. Examples of Data Management and Tracking Screens**

The online data management platform includes customized views of all modules. Under the “Participant Management” tab, the study team can obtain a summary of all individuals that have been approached, consented, and/or exited along with key demographic fields in a convenient table format. This table format allows for a personalized snapshot of all participants dependent on what is being assessed by the coordinator. For instance, in this example the participants are arranged chronologically by consent dates, with the most recent consent appearing at the top. Each participant can then be seen individually in varying “profile” views. The first icon allows the coordinator quick access to edit all participant information including contact and study information (as seen in red). The second icon prompts for a unique profile view depicting all study information, form submit dates, as well as a table of Study Tasks (as seen in purple). The “Study Tasks” table allows the coordinator to quickly assess both module completion for data entry as well as biospecimen tracking at multiple timepoints. Finally, the coordinator can view all modules/case report forms in the “Study Workflow” view (as seen in green).

**Supplementary Figure 4. Florida Pancreas Collaborative Health Screen Instrument**.

This instrument comprises a) the abridged version of the Patient-Generated Subjective Global Assessment (aPG-SGA), b) a revised version of the Edmonton Symptom Assessment System (ESAS-r), and c) the Canadian Problem Checklist.

**Supplementary Figure 5. Example of a Health Screen Report Obtained at Baseline.**

**Supplementary Figure 6. Florida Pancreas Collaborative Data Repository (FPCDR) Infrastructure**.

**Supplementary Figure 7. Total number of participants enrolled in the Florida Pancreas Collaborative, by study site and race/ethnicity**. Note: These numbers reflect enrollment through August 31, 2020.

**Supplementary Figure 8. Resources Ascertained through the Florida Pancreas Collaborative (FPC) versus the Pancreatic Ductal Adenocarcinoma (PAAD) Cohort included in The Cancer Genome Atlas (TCGA) Pan-Cancer Analyses**. This comparison reveals that the FPC: a) includes participants recruited from the community and has a higher proportion of underserved minority populations, b) collects tissue types (untreated and treated) in addition to pancreas tumors and normal pancreas, and has c) correlative blood, d) CT scans, and e) well-annotated data corresponding to each participant. Note: The PAAD cohort being referred to was published by Liu et al 2018 (*Cell* 173, 400-416).

## References

1. American Cancer Society. Cancer Facts and Figures 2020. Atlanta: American Cancer Society, 2020.

2. Rahib, L. et al. Projecting cancer incidence and deaths to 2030: the unexpected burden of thyroid, liver, and pancreas cancers in the United States. Cancer Res 74, 2913–21 (2014).

3. Howlader N, Noone AM, Krapcho M, Miller D, Bishop K, Altekruse SF, Kosary CL, Yu M, Ruhl J, Tatalovich Z, Mariotto A, Lewis DR, Chen HS, Feuer EJ, Cronin KA (eds). SEER Cancer Statistics Review, 1975-2013, National Cancer Institute. Bethesda, MD, http://seer.cancer.gov/csr/1975_2013/, based on November 2015 SEER data submission, posted to the SEER web site, April 2016.

4. Abraham, A. et al. Disparities in pancreas cancer care. Ann Surg Oncol 20, 2078–87 (2013).

5. Chang, K.J., Parasher, G., Christie, C., Largent, J. & Anton-Culver, H. Risk of pancreatic adenocarcinoma: disparity between African Americans and other race/ethnic groups. Cancer 103, 349–57 (2005).

6. Riall, T.S., Townsend, C.M., Jr., Kuo, Y.F., Freeman, J.L. & Goodwin, J.S. Dissecting racial disparities in the treatment of patients with locoregional pancreatic cancer: a 2-step process. Cancer 116, 930–9 (2010).

7. Singal, V., Singal, A.K. & Kuo, Y.F. Racial disparities in treatment for pancreatic cancer and impact on survival: a population-based analysis. J Cancer Res Clin Oncol 138, 715–22 (2012).

8. Wray, C.J., Castro-Echeverry, E., Silberfein, E.J., Ko, T.C. & Kao, L.S. A multi-institutional study of pancreatic cancer in Harris County, Texas: race predicts treatment and survival. Ann Surg Oncol 19, 2776–81 (2012).

9. Murphy, M.M. et al. Pancreatic resection: a key component to reducing racial disparities in pancreatic adenocarcinoma. Cancer 115, 3979–90 (2009).

10. Zell, J.A., Rhee, J.M., Ziogas, A., Lipkin, S.M. & Anton-Culver, H. Race, socioeconomic status, treatment, and survival time among pancreatic cancer cases in California. Cancer Epidemiol Biomarkers Prev 16, 546–52 (2007).

11. Murphy, M.M. et al. Racial differences in cancer specialist consultation, treatment, and outcomes for locoregional pancreatic adenocarcinoma. Ann Surg Oncol 16, 2968–77 (2009).

12. DeSantis, C.E. et al. Cancer statistics for African Americans, 2016: Progress and opportunities in reducing racial disparities. CA Cancer J Clin 66, 290–308 (2016).

13. American Cancer Society. Cancer Facts and Figures for African Americans 2013-2014. Atlanta: American Cancer Society, 2013.

14. American Cancer Society. Cancer Facts and Figures for Hispanics/Latinos 2012-2014. Atlanta: American Cancer Society, 2012.

15. Ahlgren, J.D. Epidemiology and risk factors in pancreatic cancer. Semin Oncol 23, 241–50 (1996).

16. (http://fcds.med.miami.edu/), F.C.D.S.

17. Permuth, J.B. et al. Racial and ethnic disparities in a state-wide registry of patients with pancreatic cancer and an exploratory investigation of cancer cachexia as a contributor to observed inequities. Cancer Med 8, 3314–3324 (2019).

18. Maisonneuve, P. & Lowenfels, A.B. Risk factors for pancreatic cancer: a summary review of meta-analytical studies. Int J Epidemiol 44, 186–98 (2015).

19. Iodice, S., Gandini, S., Maisonneuve, P. & Lowenfels, A.B. Tobacco and the risk of pancreatic cancer: a review and meta-analysis. Langenbecks Arch Surg 393, 535–45 (2008).

20. Ansary-Moghaddam, A. et al. The effect of modifiable risk factors on pancreatic cancer mortality in populations of the Asia-Pacific region. Cancer Epidemiol Biomarkers Prev 15, 2435–40 (2006).

21. Katanoda, K. et al. Population attributable fraction of mortality associated with tobacco smoking in Japan: a pooled analysis of three large-scale cohort studies. J Epidemiol 18, 251–64 (2008).

22. Lynch, S.M. et al. Cigarette smoking and pancreatic cancer: a pooled analysis from the pancreatic cancer cohort consortium. Am J Epidemiol 170, 403–13 (2009).

23. Matsuo, K. et al. Cigarette smoking and pancreas cancer risk: an evaluation based on a systematic review of epidemiologic evidence in the Japanese population. Jpn J Clin Oncol 41, 1292–302 (2011).

24. Bosetti, C. et al. Cigarette smoking and pancreatic cancer: an analysis from the International Pancreatic Cancer Case-Control Consortium (Panc4). Ann Oncol 23, 1880–8 (2012).

25. Zou, L. et al. Non-linear dose-response relationship between cigarette smoking and pancreatic cancer risk: evidence from a meta-analysis of 42 observational studies. Eur J Cancer 50, 193–203 (2014).

26. Ross, K.C. et al. Racial differences in the relationship between rate of nicotine metabolism and nicotine intake from cigarette smoking. Pharmacol Biochem Behav 148, 1–7 (2016).

27. Barreiro, E. Role of Protein Carbonylation in Skeletal Muscle Mass Loss Associated with Chronic Conditions. Proteomes 4(2016).

28. Puig-Vilanova, E. et al. Oxidative stress, redox signaling pathways, and autophagy in cachectic muscles of male patients with advanced COPD and lung cancer. Free Radic Biol Med 79, 91–108 (2015).

29. Rom, O. & Reznick, A.Z. The role of E3 ubiquitin-ligases MuRF-1 and MAFbx in loss of skeletal muscle mass. Free Radic Biol Med 98, 218–30 (2016).

30. Rom, O., Kaisari, S., Aizenbud, D. & Reznick, A.Z. Identification of possible cigarette smoke constituents responsible for muscle catabolism. J Muscle Res Cell Motil 33, 199–208 (2012).

31. Marmot, M. et al. Food, nutrition, physical activity, and the prevention of cancer: a global perspective. (2007).

32. Yuan, C. et al. Prediagnostic body mass index and pancreatic cancer survival. J Clin Oncol 31, 4229–34 (2013).

33. Coughlin, S.S., Calle, E.E., Patel, A.V. & Thun, M.J. Predictors of pancreatic cancer mortality among a large cohort of United States adults. Cancer Causes Control 11, 915–23 (2000).

34. Bracci, P.M. Obesity and pancreatic cancer: overview of epidemiologic evidence and biologic mechanisms. Mol Carcinog 51, 53–63 (2012).

35. Yeo, T.P. Demographics, epidemiology, and inheritance of pancreatic ductal adenocarcinoma. Semin Oncol 42, 8–18 (2015).

36. Chari, S.T. et al. Probability of pancreatic cancer following diabetes: a population-based study. Gastroenterology 129, 504–11 (2005).

37. Silverman, D.T. et al. Why do Black Americans have a higher risk of pancreatic cancer than White Americans? Epidemiology 14, 45–54 (2003).

38. Arnold, L.D. et al. Are racial disparities in pancreatic cancer explained by smoking and overweight/obesity? Cancer Epidemiol Biomarkers Prev 18, 2397–405 (2009).

39. Wong, S., Gu, N., Banerjee, M., Birkmeyer, J. & Birkmeyer, N. The impact of socioeconomic status on cancer care and survival. in ASCO Annual Meeting Proceedings Vol. 29 6004 (2011).

40. Sosa, J.A. et al. Importance of hospital volume in the overall management of pancreatic cancer. Ann Surg 228, 429–38 (1998).

41. Meguid, R.A., Ahuja, N. & Chang, D.C. What constitutes a “high-volume” hospital for pancreatic resection? J Am Coll Surg 206, 622 e1–9 (2008).

42. Gibney, E.M., Doshi, M.D., Hartmann, E.L., Parikh, C.R. & Garg, A.X. Health insurance status of US living kidney donors. Clin J Am Soc Nephrol 5, 912–6 (2010).

43. Noel, M. & Fiscella, K. Disparities in Pancreatic Cancer Treatment and Outcomes. Health Equity 3, 532–540 (2019).

44. Boyd, R., Lindo EG, Weeks LD, McLemore MR. On Racism: A New Standard For Publishing On Racial Health Inequities, Health Affairs Blog, July 2, 2020. DOI: 10.1377/hblog20200630.939347.

45. Halpern, M.T. Cancer disparities research: it is time to come of age. Cancer 121, 1158–9 (2015).

46. Yadav, D. et al. PROspective Evaluation of Chronic Pancreatitis for EpidEmiologic and Translational StuDies: Rationale and Study Design for PROCEED From the Consortium for the Study of Chronic Pancreatitis, Diabetes, and Pancreatic Cancer. Pancreas 47, 1229–1238 (2018).

47. Hwang, R.F. et al. Development of an integrated biospecimen bank and multidisciplinary clinical database for pancreatic cancer. Ann Surg Oncol 15, 1356–66 (2008).

48. Permuth, J.B., Trevino, J., Merchant, N. & Malafa, M. Partnering to advance early detection and prevention efforts for pancreatic cancer: the Florida Pancreas Collaborative. Future Oncol 12, 997–1000 (2016).

49. Agency for Health Care Administration. Available at http://www.fchc.state.fl.us and http://ahca.myflorida.com. Accessed August 2016.

50. Demeure, M.J. et al. Multi-institutional tumor banking: lessons learned from a pancreatic cancer biospecimen repository. Pancreas 39, 949–54 (2010).

51. Wright, D., Corner, J., Hopkinson, J. & Foster, C. Listening to the views of people affected by cancer about cancer research: an example of participatory research in setting the cancer research agenda. Health Expect 9, 3–12 (2006).

52. Permuth-Wey, J. & Borenstein, A.R. Financial Remuneration for Clinical and Behavioral Research Participation: Ethical and Practical Considerations. Ann Epidemiol (2009).

53. Lang, R. et al. African American participation in health-related research studies: indicators for effective recruitment. J Public Health Manag Pract 19, 110–8 (2013).

54. Perez, D.F., Nie, J.X., Ardern, C.I., Radhu, N. & Ritvo, P. Impact of participant incentives and direct and snowball sampling on survey response rate in an ethnically diverse community: results from a pilot study of physical activity and the built environment. J Immigr Minor Health 15, 207–14 (2013).

55. Schnieders, T., Danner, D.D., McGuire, C., Reynolds, F. & Abner, E. Incentives and barriers to research participation and brain donation among African Americans. Am J Alzheimers Dis Other Demen 28, 485–90 (2013).

56. Nishida, C., Ko, G.T. & Kumanyika, S. Body fat distribution and noncommunicable diseases in populations: overview of the 2008 WHO Expert Consultation on Waist Circumference and Waist-Hip Ratio. Eur J Clin Nutr 64, 2–5 (2010).

57. Yoshida, T. & Delafontaine, P. Mechanisms of Cachexia in Chronic Disease States. Am J Med Sci 350, 250–6 (2015).

58. Argiles, J.M., Busquets, S., Stemmler, B. & Lopez-Soriano, F.J. Cancer cachexia: understanding the molecular basis. Nat Rev Cancer 14, 754–62 (2014).

59. Barnes, A.F., Yeo, T.P., Leiby, B., Kay, A. & Winter, J.M. Pancreatic Cancer-Associated Depression: A Case Report and Review of the Literature. Pancreas 47, 1065–1077 (2018).

60. Janda, M. et al. Anxiety, depression and quality of life in people with pancreatic cancer and their carers. Pancreatology 17, 321–327 (2017).

61. Kenner, B.J. Early Detection of Pancreatic Cancer: The Role of Depression and Anxiety as a Precursor for Disease. Pancreas 47, 363–367 (2018).

62. Parker, G. & Brotchie, H. Pancreatic Cancer and Depression: A Narrative Review. J Nerv Ment Dis 205, 487–490 (2017).

63. Martin, L. et al. Prognostic factors in patients with advanced cancer: use of the patient-generated subjective global assessment in survival prediction. J Clin Oncol 28, 4376–83 (2010).

64. Howell, D., Hack, T.F., Green, E. & Fitch, M. Cancer distress screening data: translating knowledge into clinical action for a quality response. Palliat Support Care 12, 39–51 (2014).

65. Cancer Journey Portfolio. (2012). Screening for Distress, the 6th Vital Sign: A Guide to Imple-menting Best Practices in Person-Centred Care. Available at: www.cancerview.ca.

66. Roila, F. et al. Intra and interobserver variability in cancer patients’ performance status assessed according to Karnofsky and ECOG scales. Ann Oncol 2, 437–9 (1991).

67. Jones, N.C. et al. Inclusion of transgender and gender diverse health data in cancer biorepositories. Contemp Clin Trials Commun 19, 100597 (2020).

68. Land, S.R. et al. Research Priorities, Measures, and Recommendations for Assessment of Tobacco Use in Clinical Cancer Research. Clin Cancer Res 22, 1907–13 (2016).

69. Payne, T.J., Smith, P.O., McCracken, L.M., McSherry, W.C. & Antony, M.M. Assessing nicotine dependence: a comparison of the Fagerstrom Tolerance Questionnaire (FTQ) with the Fagerstrom Test for Nicotine Dependence (FTND) in a clinical sample. Addict Behav 19, 307–17 (1994).

70. Pomerleau, C.S., Pomerleau, O.F., Majchrzak, M.J., Kloska, D.D. & Malakuti, R. Relationship between nicotine tolerance questionnaire scores and plasma cotinine. Addict Behav 15, 73–80 (1990).

71. Yasmin-Karim, S. et al. Enhancing the Therapeutic Efficacy of Cancer Treatment With Cannabinoids. Front Oncol 8, 114 (2018).

72. Slavich, G.M. & Shields, G.S. Assessing Lifetime Stress Exposure Using the Stress and Adversity Inventory for Adults (Adult STRAIN): An Overview and Initial Validation. Psychosom Med 80, 17–27 (2018).

73. Buysse, D.J., Reynolds, C.F., 3rd, Monk, T.H., Berman, S.R. & Kupfer, D.J. The Pittsburgh Sleep Quality Index: a new instrument for psychiatric practice and research. Psychiatry Res 28, 193–213 (1989).

74. Schmidt, U., Simunec, D., Piso, P., Klempnauer, J. & Schlitt, H.J. Quality of life and functional long-term outcome after partial pancreatoduodenectomy: pancreatogastrostomy versus pancreatojejunostomy. Ann Surg Oncol 12, 467–72 (2005).

75. Fitzsimmons, D. et al. Development of a disease specific quality of life (QoL) questionnaire module to supplement the EORTC core cancer QoL questionnaire, the QLQ-C30 in patients with pancreatic cancer. EORTC Study Group on Quality of Life. Eur J Cancer 35, 939–41 (1999).

76. Fitzsimmons, D. et al. Symptoms and quality of life in chronic pancreatitis assessed by structured interview and the EORTC QLQ-C30 and QLQ-PAN26. Am J Gastroenterol 100, 918–26 (2005).

77. Al-Hawary, M.M. et al. Pancreatic ductal adenocarcinoma radiology reporting template: consensus statement of the society of abdominal radiology and the american pancreatic association. Gastroenterology 146, 291–304.e1 (2014).

78. National Cancer Institute Best Practices for Biospecimen Resources: National Cancer Institute, N.I.o.H., U.S. Department of Health and Human Services. (June 2007).

79. Ivanics, T. et al. Patient-derived xenograft cryopreservation and reanimation outcomes are dependent on cryoprotectant type. Lab Invest 98, 947–956 (2018).

80. Roife, D. et al. Ex Vivo Testing of Patient-Derived Xenografts Mirrors the Clinical Outcome of Patients with Pancreatic Ductal Adenocarcinoma. Clin Cancer Res 22, 6021–6030 (2016).

81. Roife, D. et al. Generation of patient-derived xenografts from fine needle aspirates or core needle biopsy. Surgery 161, 1246–1254 (2017).

82. Delitto, D. et al. Human pancreatic cancer xenografts recapitulate key aspects of cancer cachexia. Oncotarget (2016).

83. Go, K.L. et al. Orthotopic Patient-Derived Pancreatic Cancer Xenografts Engraft Into the Pancreatic Parenchyma, Metastasize, and Induce Muscle Wasting to Recapitulate the Human Disease. Pancreas (2017).

84. Quantitative Imaging Network. National Cancer Institute. Available from: http://imaging.cancer.gov/informatics/qin; [Aug 2016].

85. Clark, K. et al. The Cancer Imaging Archive (TCIA): maintaining and operating a public information repository. J Digit Imaging 26, 1045–57 (2013).

86. Al-Hawary, M.M. et al. Pancreatic ductal adenocarcinoma radiology reporting template: consensus statement of the society of abdominal radiology and the american pancreatic association. Gastroenterology 146, 291–304 e1 (2014).

87. Vigano, A., Del Fabbro, E., Bruera, E. & Borod, M. The cachexia clinic: from staging to managing nutritional and functional problems in advanced cancer patients. Crit Rev Oncog 17, 293–303 (2012).

88. Oldenmenger, W.H., de Raaf, P.J., de Klerk, C. & van der Rijt, C.C. Cut points on 0-10 numeric rating scales for symptoms included in the Edmonton Symptom Assessment Scale in cancer patients: a systematic review. J Pain Symptom Manage 45, 1083–93 (2013).

89. Chang, J.I., Huang, B.Z. & Wu, B.U. Impact of Integrated Health Care Delivery on Racial and Ethnic Disparities in Pancreatic Cancer. Pancreas 47, 221–226 (2018).

90. Luque, J.S. et al. Formative research on perceptions of biobanking: what community members think. J Cancer Educ 27, 91–9 (2012).

91. Schmotzer, G.L. Barriers and facilitators to participation of minorities in clinical trials. Ethn Dis 22, 226–30 (2012).

92. Sheppard, V.B., Mays, D., LaVeist, T. & Tercyak, K.P. Medical mistrust influences black women’s level of engagement in BRCA 1/2 genetic counseling and testing. J Natl Med Assoc 105, 17–22 (2013).

93. Halbert, C.H., McDonald, J., Vadaparampil, S., Rice, L. & Jefferson, M. Conducting Precision Medicine Research with African Americans. PLoS One 11, e0154850 (2016).

94. Reddy, A. et al. Privacy Concerns About Personal Health Information and Fear of Unintended Use of Biospecimens Impact Donations by African American Patients. J Cancer Educ 35, 522–529 (2020).

95. Davis, T.C., Arnold, C.L., Mills, G. & Miele, L. A Qualitative Study Exploring Barriers and Facilitators of Enrolling Underrepresented Populations in Clinical Trials and Biobanking. Front Cell Dev Biol 7, 74 (2019).

96. Joseph, G. & Dohan, D. Recruiting minorities where they receive care: Institutional barriers to cancer clinical trials recruitment in a safety-net hospital. Contemp Clin Trials 30, 552–9 (2009).

97. Cook, E.D. et al. Recruitment practices for U.S. minority and underserved populations in NRG oncology: Results of an online survey. Contemp Clin Trials Commun 10, 100–104 (2018).

98. Bailey, P. et al. Genomic analyses identify molecular subtypes of pancreatic cancer. Nature 531, 47–52 (2016).

99. Collisson, E.A. et al. Subtypes of pancreatic ductal adenocarcinoma and their differing responses to therapy. Nat Med 17, 500–3 (2011).

100. Moffitt, R.A. et al. Virtual microdissection identifies distinct tumor- and stroma-specific subtypes of pancreatic ductal adenocarcinoma. Nat Genet 47, 1168–78 (2015).

101. Integrated Genomic Characterization of Pancreatic Ductal Adenocarcinoma. Cancer Cell 32, 185–203.e13 (2017).

102. Liu, J. et al. An Integrated TCGA Pan-Cancer Clinical Data Resource to Drive High-Quality Survival Outcome Analytics. Cell 173, 400–416.e11 (2018).

103. Kiviniemi, M.T. et al. Pilot intervention outcomes of an educational program for biospecimen research participation. J Cancer Educ 28, 52–9 (2013).

104. Adams-Campbell, L.L. et al. Predictors of biospecimen donation in the Black Women’s Health Study. Cancer Causes Control 27, 797–803 (2016).

105. Beato F R.D., Dezsi KB, Ortiz A, Johnson JO, Chen DT, Ali K, Yoder SJ, Jeong D, Malafa M, Hodul P, Jiang K, Centeno BA, Abdallah MA, Balasi JA, Tassielli AF, Sarcar B, Teer JK, DeNicola GM, Permuth JB, Fleming JB. Establishing a LivingBiobank of Patient-Derived Organoids of Intraductal Papillary Mucinous Neoplasms of the Pancreas. bioRxiv doi: https://doi.org/10.1101/2020.09.11.283168.

