## Supplementary material for "The Florida Pancreas Collaborative Next-Generation Biobank: State-wide Infrastructure to Reduce Disparities and Improve Survival for a Racially and Ethnically Diverse Cohort of Patients with Pancreatic Cancer": Supp Fig 1-8

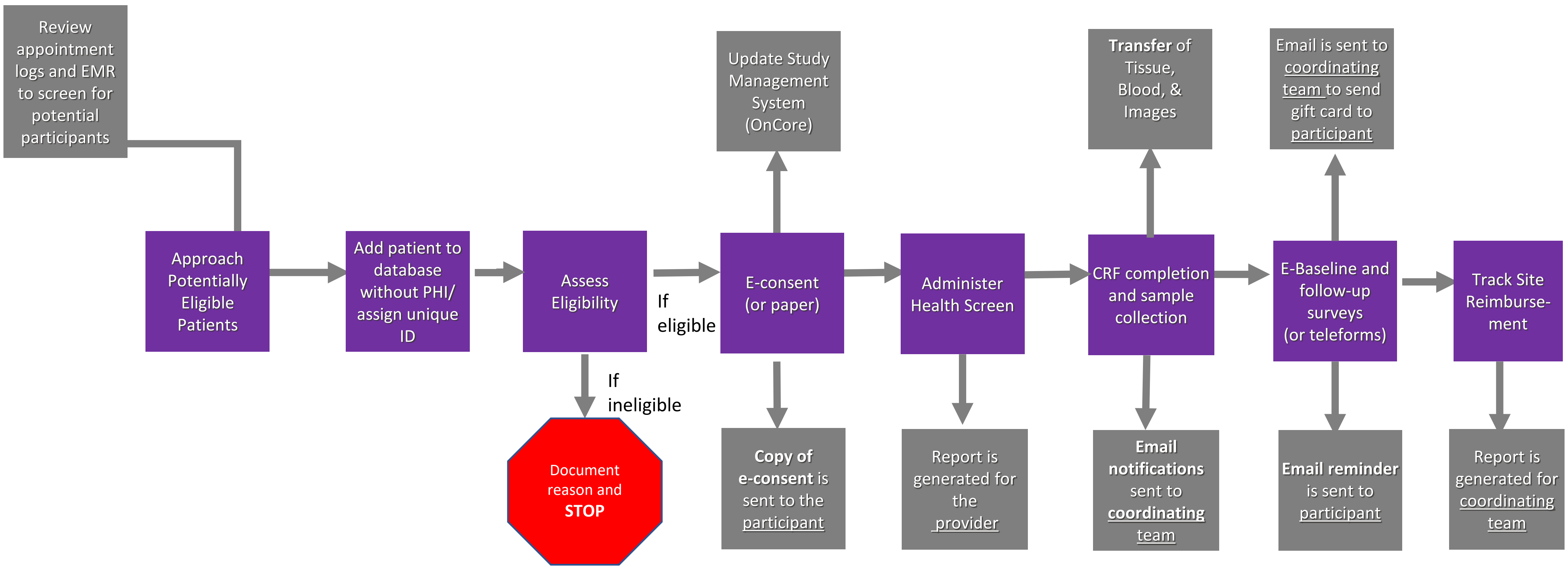
Review appointment logs and EMR to screen for potential participants
Approach Potentially Eligible Patients
Add patient to database without PHI/ assign unique ID
Assess Eligibility
If ineligible
Document reason and STOP
If eligible
E-consent (or paper)
Update Study Management System (OnCore)
Copy of e-consent is sent to the participant
Administer Health Screen
Report is generated for the provider
CRF completion and sample collection
Transfer of Tissue, Blood, & Images
Email notifications sent to coordinating team
E-Baseline and follow-up surveys (or teleforms)
Email is sent to coordinating team to send gift card to participant
Email reminder is sent to participant
Track Site Reimburse-ment
Report is generated for coordinating team

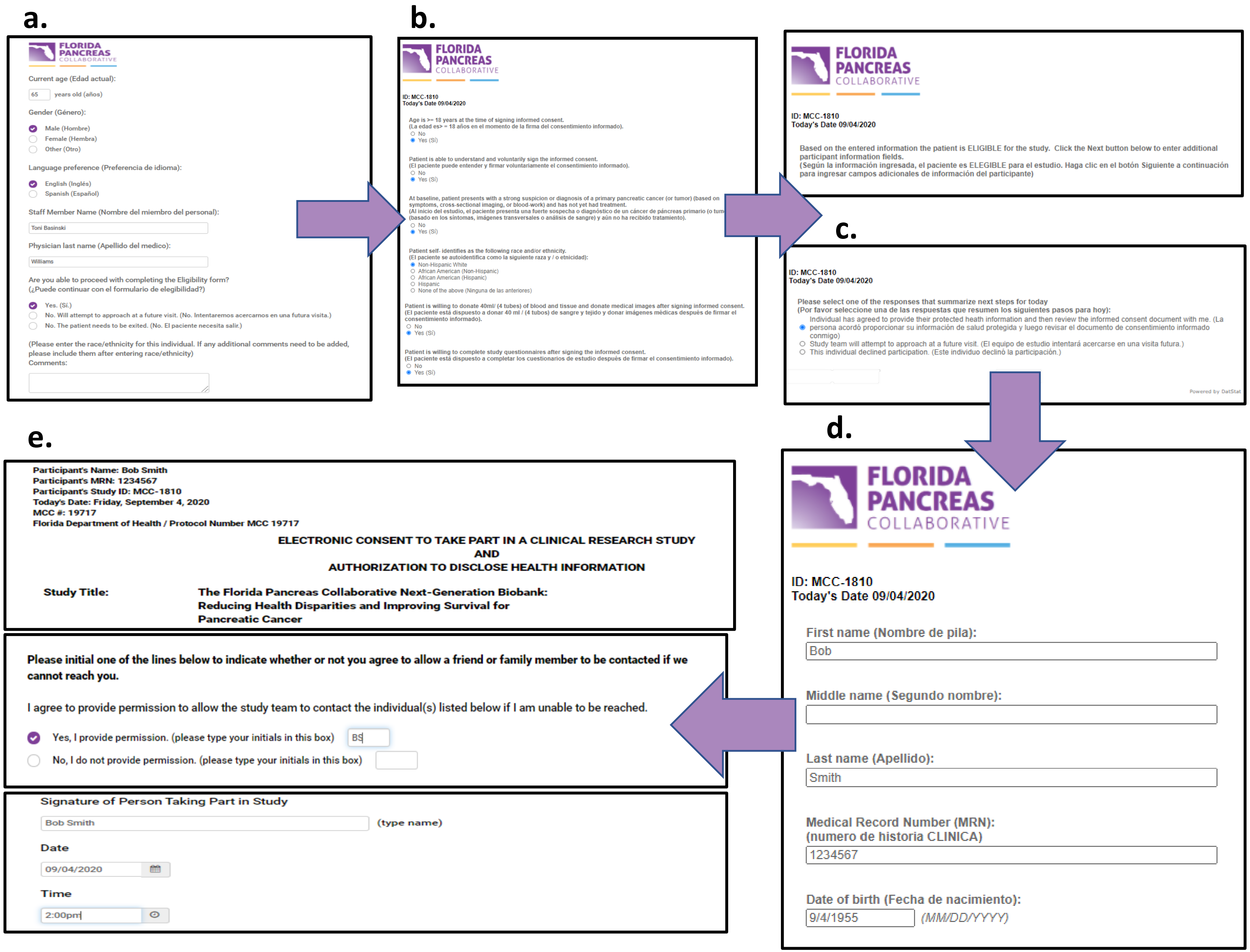
a.
FLORIDA PANCREAS COLLABORATIVE
Current age (Edad actual):
65 years old (años)
Gender (Género):
Male (Hombre)
Female (Hembra)
Other (Otro)
Language preference (Preferencia de idioma):
English (Inglés)
Spanish (Español)
Staff Member Name (Nombre del miembro del personal):
Toni Basinski
Physician last name (Apellido del medico):
Williams
Are you able to proceed with completing the Eligibility form?
(¿Puede continuar con el formulario de elegibilidad?)
Yes. (Sí.)
No. Will attempt to approach at a future visit. (No. Intentaremos acercarnos en una futura visita.)
No. The patient needs to be exited. (No. El paciente necesita salir.)
(Please enter the race/ethnicity for this individual. If any additional comments need to be added, please include them after entering race/ethnicity)
Comments:
b.
FLORIDA PANCREAS COLLABORATIVE
ID: MCC-1810
Today's Date 09/04/2020
Age is >= 18 years at the time of signing informed consent.
No
Yes (Sí)
Patient is able to understand and voluntarily sign the informed consent.
No
Yes (Sí)
No
Yes (Sí)
Patient self- identifies as the following race and/or ethnicity.
Non-Hispanic White
African American (Non-Hispanic)
African American (Hispanic)
Hispanic
None of the above (Ninguna de las anteriores)
No
Yes (Sí)
Patient is willing to complete study questionnaires after signing the informed consent.
No
Yes (Sí)
FLORIDA PANCREAS COLLABORATIVE
ID: MCC-1810
Today's Date 09/04/2020
Based on the entered information the patient is ELIGIBLE for the study. Click the Next button below to enter additional participant information fields.
(Según la información ingresada, el paciente es ELEGIBLE para el estudio. Haga clic en el botón Siguiente a continuación para ingresar campos adicionales de información del participante)
c.
ID: MCC-1810
Today's Date 09/04/2020
Please select one of the responses that summarize next steps for today
(Por favor seleccione una de las respuestas que resumen los siguientes pasos para hoy):
Study team will attempt to approach at a future visit. (El equipo de estudio intentará acercarse en una visita futura.)
This individual declined participation. (Este individuo declinó la participación.)
Powered by DatStat
d.
FLORIDA PANCREAS COLLABORATIVE
ID: MCC-1810
Today's Date 09/04/2020
First name (Nombre de pila):
Bob
Middle name (Segundo nombre):
Last name (Apellido):
Smith
Medical Record Number (MRN):
(numero de historia CLINICA)
1234567
Date of birth (Fecha de nacimiento):
9/4/1955
(MM/DD/YYYY)
e.
Participant's Name: Bob Smith
Participant's MRN: 1234567
Participant's Study ID: MCC-1810
Today's Date: Friday, September 4, 2020
MCC #: 19717
Florida Department of Health / Protocol Number MCC 19717
ELECTRONIC CONSENT TO TAKE PART IN A CLINICAL RESEARCH STUDY
AND
AUTHORIZATION TO DISCLOSE HEALTH INFORMATION
Study Title:
The Florida Pancreas Collaborative Next-Generation Biobank: Reducing Health Disparities and Improving Survival for Pancreatic Cancer
Please initial one of the lines below to indicate whether or not you agree to allow a friend or family member to be contacted if we cannot reach you.
I agree to provide permission to allow the study team to contact the individual(s) listed below if I am unable to be reached.
Yes, I provide permission. (please type your initials in this box)
BS
No, I do not provide permission. (please type your initials in this box)
Signature of Person Taking Part in Study
Bob Smith
(type name)
Date
09/04/2020
Time
2:00pm

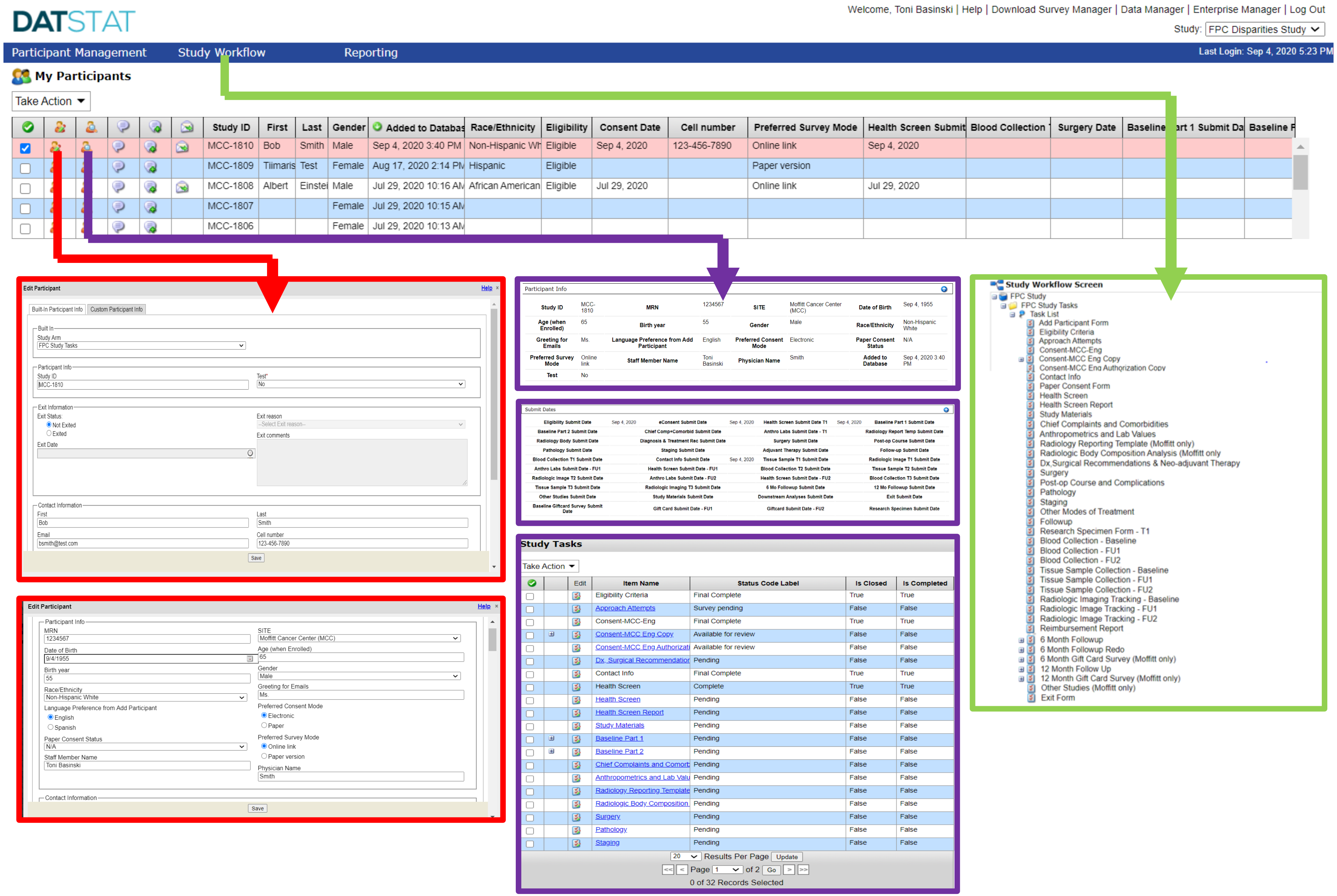
DATSTAT
Welcome, Toni Basinski | Help | Download Survey Manager | Data Manager | Enterprise Manager | Log Out
Study: FPC Disparities Study
Participant Management
Study Workflow
Reporting
Last Login: Sep 4, 2020 5:23 PM
My Participants
Take Action
Study ID
First
Last
Gender
Added to Databas
Race/Ethnicity
Eligibility
Consent Date
Cell number
Preferred Survey Mode
Health Screen Submit
Blood Collection
Surgery Date
Baseline
Baseline F
MCC-1810 Bob Smith Male Sep 4, 2020 3:40 PM Non-Hispanic Wh Eligible Sep 4, 2020 123-456-7890 Online link Sep 4, 2020
MCC-1809 Tiimaris Test Female Aug 17, 2020 2:14 PM Hispanic Eligible Paper version
MCC-1808 Albert Einstei Male Jul 29, 2020 10:16 AM African American Eligible Jul 29, 2020 Online link Jul 29, 2020
MCC-1807 Female Jul 29, 2020 10:15 AM
MCC-1806 Female Jul 29, 2020 10:13 AM
Edit Participant
Built-In Participant Info
Custom Participant Info
Study Arm
FPC Study Tasks
Participant Info
Study ID
MCC-1810
Exit Information
Exit Status
Not Exited
Exited
Exit Date
Exit reason
Exit comments
Contact Information
First
Bob
Last
Smith
Email

Cell number
123-456-7890
Save
Edit Participant
MRN
1234567
SITE
Moffitt Cancer Center (MCC)
Date of Birth
9/4/1955
Age (when Enrolled)
65
Birth year
55
Gender
Male
Race/Ethnicity
Non-Hispanic White
Greeting for Emails
Ms.
Language Preference from Add Participant
English
Spanish
Preferred Consent Mode
Electronic
Paper
Paper Consent Status
N/A
Preferred Survey Mode
Online link
Paper version
Staff Member Name
Toni Basinski
Physician Name
Smith
Save
Participant Info
Submit Dates
Study Tasks
Take Action
Edit
Item Name
Status Code Label
Is Closed
Is Completed
Eligibility Criteria Final Complete True True
Approach Attempts Survey pending False False
Consent-MCC-Eng Final Complete True True
Consent-MCC Eng Copy Available for review False False
Health Screen Complete True True
Study Materials Pending False False
Baseline Part 1 Pending False False
Baseline Part 2 Pending False False
Surgery Pending False False
Pathology Pending False False
Staging Pending False False
Results Per Page
Update
Page 1 of 2
Go
0 of 32 Records Selected
Study Workflow Screen
FPC Study
FPC Study Tasks
Task List
Add Participant Form
Eligibility Criteria
Approach Attempts
Consent-MCC-Eng
Consent-MCC Eng Copy
Contact Info
Paper Consent Form
Health Screen
Health Screen Report
Study Materials
Chief Complaints and Comorbidities
Anthropometrics and Lab Values
Radiology Reporting Template (Moffitt only)
Surgery
Post-op Course and Complications
Pathology
Staging
Other Modes of Treatment
Followup
Research Specimen Form - T1
Blood Collection - Baseline
Blood Collection - FU1
Blood Collection - FU2
Tissue Sample Collection - Baseline
Tissue Sample Collection - FU1
Tissue Sample Collection - FU2
Radiologic Imaging Tracking - Baseline
Radiologic Image Tracking - FU1
Radiologic Image Tracking - FU2
Reimbursement Report
6 Month Followup
6 Month Followup Redo
6 Month Gift Card Survey (Moffitt only)
12 Month Follow Up
12 Month Gift Card Survey (Moffitt only)
Other Studies (Moffitt only)
Exit Form

**a.**

Patient ID Information

☐☐☐ - ☐☐☐☐

Draft

History (Boxes 1-4 are designed to be completed by the patient.)

**1. Weight:** *(See Worksheet 1)*

In summary of my current and recent weight:

I currently weigh about ☐☐☐ pounds

I am about ☐ feet ☐☐ tall

One month ago I weighed about ☐☐☐ pounds

Six months ago I weighed about ☐☐☐ pounds

During the past two weeks my weight has:

☐ decreased (1) ☐ not changed (0) ☐ increased (0)

Box 1 ☐

**2. Food Intake:** As compared to my normal intake, I would rate my food intake during the past month as:

☐ unchanged (0)
☐ more than usual (0)
☐ less than usual (1)

I am now taking:

☐ *normal food* but less than normal amount (1)
☐ little solid food (2)
☐ only liquids (3)
☐ only nutritional supplements (3)
☐ very little of anything (4)
☐ only tube feedings or only nutrition by vein (0)

Box 2 ☐

**3. Symptoms:** I have had the following problems that have kept me from eating enough during the past two weeks (check all that apply):

☐ no problems eating (0)
☐ no appetite, just did not feel like eating (0)
☐ nausea (1)
☐ constipation (1)
☐ mouth sores (2)
☐ things taste funny or have no taste (1)
☐ problems swallowing (2)
☐ pain; where? (3) ____________
☐ other** (1) ____________
☐ vomiting (3)
☐ diarrhea (3)
☐ dry mouth (1)
☐ smells bother me (1)
☐ feel full quickly (1)
☐ fatigue (1)

** Examples: depression, money, or dental problems

Box 3 ☐

**4. Activities and Function:** Over the past month, I would generally rate my activity as:

☐ normal with no limitations (0)
☐ not my normal self, but able to be up and about with fairly normal activities (1)
☐ not feeling up to most things, but in bed or chair less than half the day (2)
☐ able to do little activity and spend most of the day in bed or chair (3)
☐ pretty much bedridden, rarely out of bed (3)

Box 4 ☐

©FD Ottery 2005, 2006, 2015 v3.22.15

**Additive Score of the Boxes 1-4** ☐☐



**b.**

Please select the number that best describes how you feel NOW:

| | None 0 | 1 | 2 | 3 | 4 | 5 | 6 | 7 | 8 | 9 | Worst Possible 10 |
|---|---|---|---|---|---|---|---|---|---|---|---|
| Pain | ☐ | ☐ | ☐ | ☐ | ☐ | ☐ | ☐ | ☐ | ☐ | ☐ | ☐ |
| Tiredness | ☐ | ☐ | ☐ | ☐ | ☐ | ☐ | ☐ | ☐ | ☐ | ☐ | ☐ |
| Drowsiness | ☐ | ☐ | ☐ | ☐ | ☐ | ☐ | ☐ | ☐ | ☐ | ☐ | ☐ |
| Nausea | ☐ | ☐ | ☐ | ☐ | ☐ | ☐ | ☐ | ☐ | ☐ | ☐ | ☐ |
| Lack of appetite | ☐ | ☐ | ☐ | ☐ | ☐ | ☐ | ☐ | ☐ | ☐ | ☐ | ☐ |
| Shortness of Breath | ☐ | ☐ | ☐ | ☐ | ☐ | ☐ | ☐ | ☐ | ☐ | ☐ | ☐ |
| Depression *(Depression=feeling sad)* | ☐ | ☐ | ☐ | ☐ | ☐ | ☐ | ☐ | ☐ | ☐ | ☐ | ☐ |
| Anxiety *(Anxiety=feeling nervous)* | ☐ | ☐ | ☐ | ☐ | ☐ | ☐ | ☐ | ☐ | ☐ | ☐ | ☐ |
| Best Wellbeing *(Wellbeing=how you feel overall)* | ☐ | ☐ | ☐ | ☐ | ☐ | ☐ | ☐ | ☐ | ☐ | ☐ | ☐ |
| Other Problem *(for example constipation):* | ☐ | ☐ | ☐ | ☐ | ☐ | ☐ | ☐ | ☐ | ☐ | ☐ | ☐ |

**c.**

Draft

Please check all of the following items that have been a concern or problem for you in the past week including today:

**Emotional:**

☐ Fears/Worries
☐ Sadness
☐ Frustration/Anger
☐ Changes in appearance
☐ Intimacy/Sexuality
☐ Change in who I am

**Spiritual:**

☐ Meaning/Purpose of life
☐ Faith

**Practical:**

☐ Work/School
☐ Finances
☐ Getting to and from appointments
☐ Accommodation
☐ Quitting Smoking

**Social/Family:**

☐ Feeling a burden to otherss
☐ Worry about family/friends
☐ Feeling alone

**Informational:**

☐ Understanding my illness and/or treatment
☐ Talking with the health care team
☐ Making treatment decisions
☐ Knowing about available resources
☐ Taking medications as prescribed

**Physical:**

☐ Concentration/Memory
☐ Sleep
☐ Weight

Have you smoked at least 100 cigarettes in your lifetime?

☐ No ☐ Yes

Have you smoked any cigarettes in the past 30 days?

☐ No ☐ Yes

FLORIDA PANCREAS COLLABORATIVE

Wed Sep 9 2020

Dear Dr. Fleming and the clinical research team at MCC,

Below please find a report that summarizes survey responses provided by your patient, John Smith. His survey responses were screened for several conditions (cancer cachexia, depression, and distress) and an exposure (tobacco use) that can adversely impact an oncology patient's quality of life. **Based on our preliminary assessment, it appears that Mr. Smith may have or be at risk for Refractory Cachexia, high/severe risk of depression, and distress. Mr. Smith also reports regular tobacco use.**

***Concern 1: Mr. Smith may have or be at risk for <u>Refractory Cachexia</u> because he reported:***

- 20% weight loss over the past 6 months
- A poor appetite
- *his food intake is less than usual. He is now taking: little solid food*
- *Symptoms that have kept him from eating enough in the past two weeks, including:*
  - ***no appetite, just did not feel like eating***
  - ***constipation***
  - ***mouth sores***
  - ***things taste funny or have no taste***
  - ***problems swallowing***
- *Lower level of activities or functioning over the last month such as: not feeling up to most things, but in bed or chair less than half the day*

***Recommendation 1: Consider referring Mr. Smith for palliative care, psychosocial support, and/or a discussion regarding nutritional support.***

***Concern 2: Mr. Smith may have or be at risk for <u>severe depression</u> because he reported:***

- A value of 10 on a scale of 1 (no depression) to 10 (worst possible depression)

***Recommendation 2: Consider referring Mr. Smith to an appropriate professional (ie. a psychiatrist) for definitive diagnosis. Non-pharmacological (patient education and information, counseling, psychotherapy, behavioral support) or pharmacological agents (selective serotonin reuptake inhibitors (SSRIs) and tricyclic antidepressants) may be indicated.***

***Concern 3: Mr. Smith may have or be at risk for <u>distress</u> due to symptoms and/or concerns that may be emotional, practical, informational, spiritual, or physical in nature. He reported:***

- 8 for Pain
- 9 for Tiredness
- 8 for Drowsiness
- 7 for Nausea
- 5 for Shortness of Breath
- 10 for Anxiety (Anxiety=feeling nervous)
- 9 for Best Well being (Well being=how you feel overall)
- Fears/Worries
- Sadness
- Frustration/Anger
- Getting to and from appointments
- Quitting smoking
- Understanding my illness and/or treatment
- Knowing about available resources
- Feeling a burden to others
- Worry about family/friends
- Feeling alone
- Sleep
- Weight

***Recommendation 3: Consider referring Mr. Smith to an appropriate professional for counseling (ie. a social worker) and/or to other services (ie. a chaplain).***

***Concern 4: Mr. Smith reports that he has smoked cigarettes in the last 30 days.***

***Recommendation 4: Advise Mr. Smith*** **that quitting smoking is the most important thing he can do to protect his health now and in the future. Continuing to smoke may make his cancer and/or cancer treatment worse. Quitting may dramatically improve his recovery and quality of life. Cutting down while he is ill is not enough as occasional or light smoking is still dangerous.**

**Please provide him with a brochure that contains information that can help him take steps to quit smoking. The brochure also includes information regarding** *the Florida Department of Health's Tobacco Free Florida Quitline at 877-U-CAN-NOW* **that provides telephone counseling services and a website where he can find smoking cessation support in different modalities (e.g., group therapy, online, self-help booklets).**

If you have questions, please contact the study team at or 800-456-3434 x4715

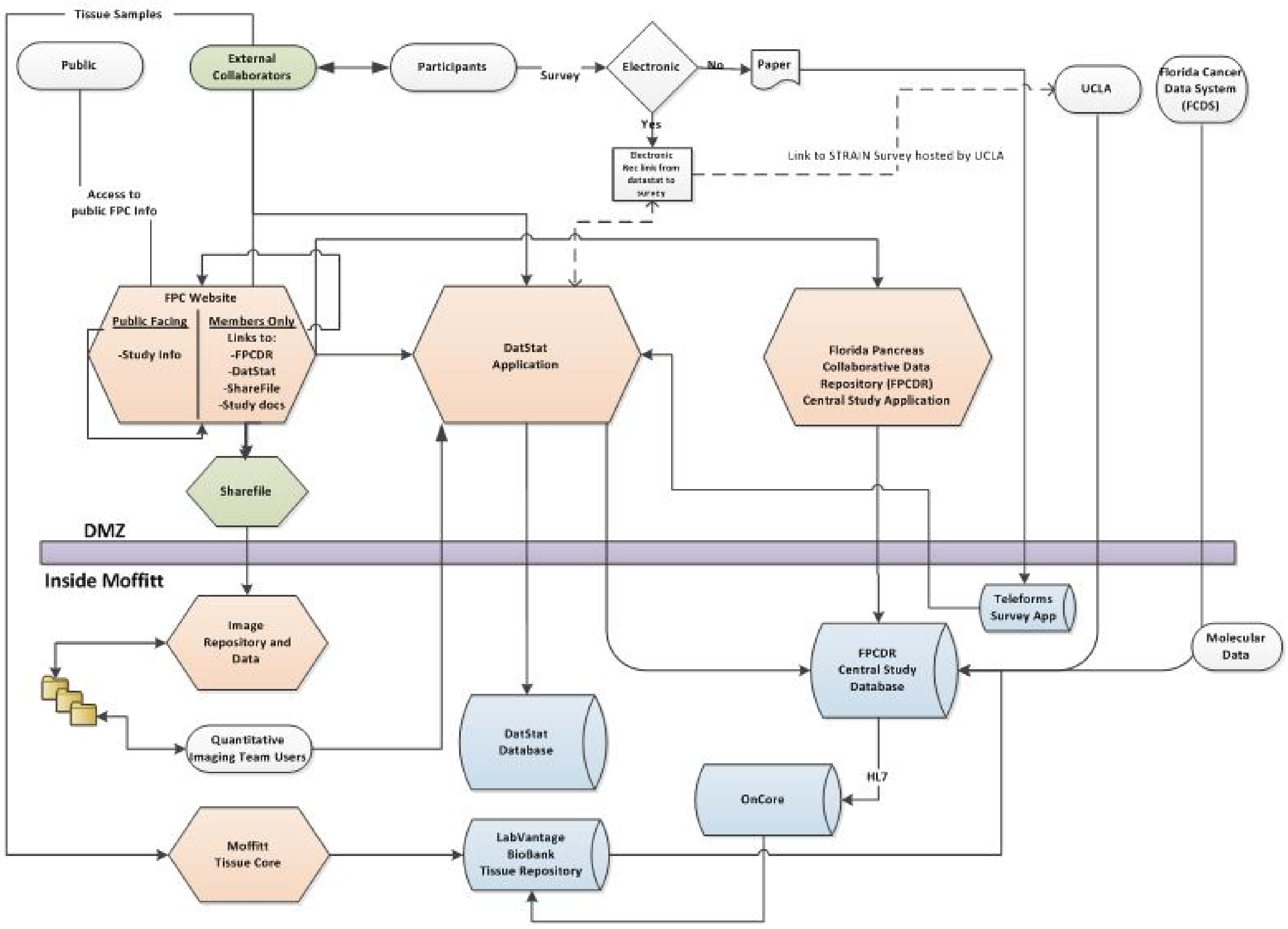

Tissue Samples
Public
External Collaborators
Participants
Survey
Electronic
No
Paper
UCLA
Florida Cancer Data System (FCDS)
Yes
Electronic Rec link from datastat to survey
Link to STRAIN Survey hosted by UCLA
Access to public FPC Info
FPC Website
Public Facing
-Study Info
Members Only
Links to:
-FPCDR
-DatStat
-ShareFile
-Study docs
DatStat Application
Florida Pancreas Collaborative Data Repository (FPCDR) Central Study Application
Sharefile
DMZ
Inside Moffitt
Teleforms Survey App
Image Repository and Data
FPCDR Central Study Database
Molecular Data
Quantitative Imaging Team Users
DatStat Database
HL7
OnCore
Moffitt Tissue Core
LabVantage BioBank Tissue Repository

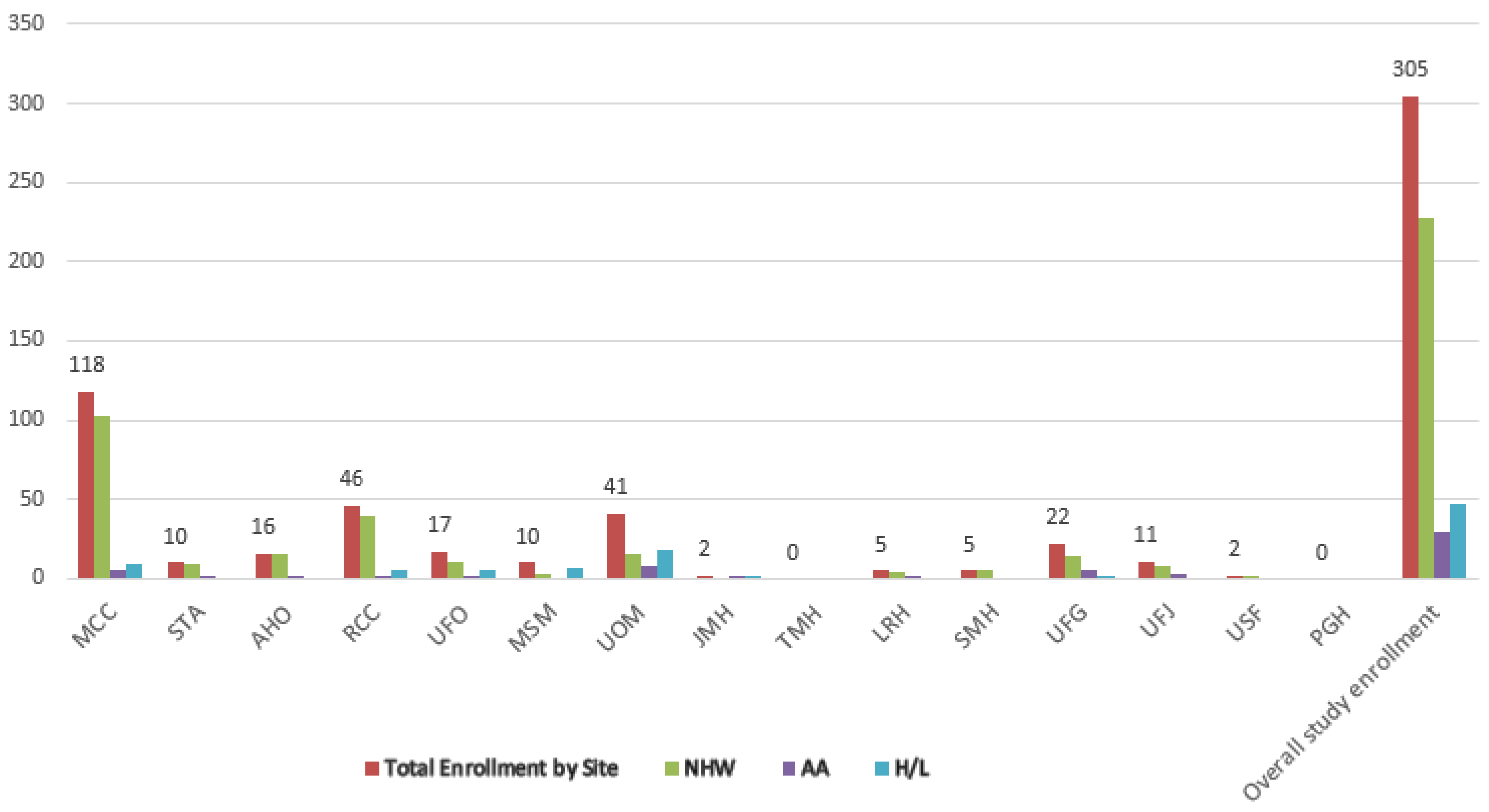
350
300
250
200
150
100
50
0
118
10
16
46
17
10
41
2
0
5
5
22
11
2
0
305
MCC
STA
AHO
RCC
UFO
MSM
UOM
JMH
TMH
LRH
SMH
UFG
UFJ
USF
PGH
Overall study enrollment
Total Enrollment by Site
NHW
AA
H/L

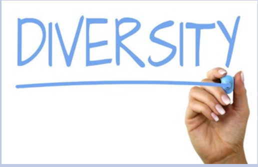


**Diverse Population of Pancreatic Cases from Academic Cancer Centers & Community Hospitals.**

- AA and H/L will each comprise at least 10-15% of the FPC study population.
- *The TCGA PAAD cohort primarily derives from major cancer centers; AA and H/L each represent <5% of the population.*

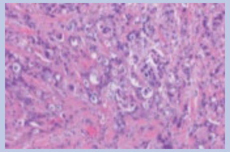

**Multiple Tissue Types Are Collected Pre- or Post- Treatment.**

- FPC collects pancreas tumor, normal pancreas, subcutaneous & omental adipose, & muscle tissues.
- *The TCGA PAAD cohort includes untreated pancreas tumor and normal pancreas tissue.*

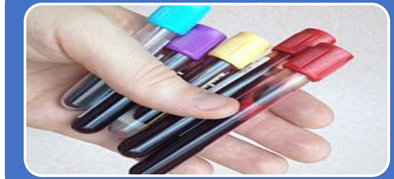

**Blood is collected*.**

- FPC processes for whole blood, plasma, and serum.
- *The TCGA PAAD cohort does not mention correlative blood products for ancestry informative analyses or circulating biomarker analyses.*

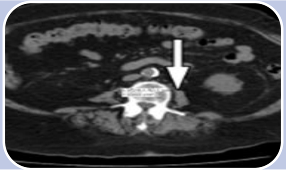

**Standard-of-Care Computed Tomography (CT) Scans Are Collected*.**

- FPC is acquiring and using CTs to evaluate radiographic criteria and for body composition analyses.
- *The TCGA PAAD cohort does not showcase correlative CT images.*

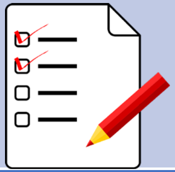

**Well annotated clinical, epidemiologic, laboratory, and quality of life data*.**

- FPC is collecting data from self-administered questionnaires, the electronic medical record, and cancer registry.
- *The TCGA PAAD cohort is extremely limited in clinical annotation.*

*at baseline, 6 months, and 12 months
