## Supplementary material for "The Florida Pancreas Collaborative Next-Generation Biobank: State-wide Infrastructure to Reduce Disparities and Improve Survival for a Racially and Ethnically Diverse Cohort of Patients with Pancreatic Cancer": Supp Table 1

**Supplementary Table 1. Validated Instruments Incoprorated into the FPC Health Screen or Comprehensive Questionnaire.**

| Survey Name | Abbreviation | Where Administered | Purpose of Survey |
|---|---|---|---|
| Edmonton Symptom Assessment Scale | ESAS | Health Screen | To assess nine commonly observed symptoms in cancer patients i.e. pain, tiredness, nausea, depression, anxiety, drowsiness, appetite, wellbeing and shortness of breath and determine the clinical profile of the symptoms over time. |
| Patient Generated - Subjective Global Assessment Short form | PG-SGA | Health Screen | PG-SGA consists of four main sections, ie. Weight, Food Intake, Symptoms and Activities that helps to determine the functional status of the patient. |
| Pittsburgh Sleep Quality Index | PSQI | Questionnaire | In PSQI, using the 19 individual items, 7 "component scores" are generated, assessing sleep quality, sleep latency, duration, habitual sleep efficiency, sleep disturbances, use of sleep medications and daytime dysfunction. Information is collected for the past one month. |
| Cancer Patient Tobacco Use Questionnaire | C-TUQ | Questionnaire | NCI AACR Cancer Patient Tobacco Use Assessment Task Force developed and validated the C-TUQ. The survey collects information on smoking status, smoking history and status relative to cancer diagnosis and treatment, use of tobacco products and secondhand smoke exposure and cessation. |
| European Organization for Research and Treatment of Cancer – Quality of Life of Cancer Patients | EORTC QLQ-C3( | Questionnaire | QLQ C30 is a cancer-specific quality of life questionnaire consisting of five functional scales, three symptom scales, an overall health status and commonly reported symptoms by cancer patients and perceived financial effect of the disease. |
| European Organization for Research and Treatment of Cancer – Pancreatic Cancer (in phase III of testing) | EORTC PAN26 | Questionnaire | QLQ-PAN26 consists of 26 four level likert scale questions focussing on pancreatic pain scale referring to abdominal discomfort, back pain, pain during night and discomfort in certain positions. |
| Enhancing Recovery in Coronary Heart Disease Social Support Inventory | ENRICHD-ESSI | Questionnaire | ESSI is a seven item survey measuring the range of social support in the patients life using a Likert scale for the first 6 questions. |
| Life Orientation Test – Revised | LOTR | Questionnaire | LOT-R includes 10 questions and helps in determining the individual differences in generalized optimism versus pessimism. The revised version also adds more details on expections for the future. |
| Stress and Adversity Inventory | STRAIN | Questionnaire | A stress assessment tool available online and evaluating the patient's exposure to acute and chronic stress throughout their lifetime. |
| Dietary Screener Questionnaire | DSQ | Questionnaire | DSQ includes dietary factors that are of interest in cancer and heart disease and collects dietary intake over the past month. |
