## Supplementary material for "The Florida Pancreas Collaborative Next-Generation Biobank: State-wide Infrastructure to Reduce Disparities and Improve Survival for a Racially and Ethnically Diverse Cohort of Patients with Pancreatic Cancer": Supp Table 2

| Supplementary Table 2. Sensitivity analysis comparing demographic and clinical characteristics by follow-up status. | | | | |
|---|---|---|---|---|
| | | Exited | | |
| **Variable** | **All participants (n=305)** | **No (n=258)** | **Yes (n=47)** | ***P*-Value** |
| Age (years), mean (+/- SD) | 68 (10.6) | 68 (10.6) | 70 (10.5) | 0.3190 |
| Gender, n (%) | | | | |
| Female | 161 (52.8) | 136 (52.7) | 25 (53.2) | 0.9518 |
| Male | 144 (47.2) | 122 (47.3) | 22 (46.8) | |
| Race/Ethnicity†, n (%) | | | | |
| African American (AA) | 30 (9.8) | 27 (10.5) | 3 (6.4) | 0.4741 |
| Hispanic/Latinx (H/L) | 47 (15.4) | 42 (16.3) | 5 (10.6) | |
| Non-Hispanic White (NHW) | 228 (74.8) | 189 (73.3) | 39 (83.0) | |
| Education Level†, n (%) | | | | |
| High school or GED | 46 (31.1) | 39 (31.7) | 7 (28.0) | 0.9047 |
| College | 65 (43.9) | 54 (43.9) | 11 (44.0) | |
| Post graduate | 37 (25.0) | 30 (24.4) | 7 (28.0) | |
| Data not yet availableᶲ | 157 | 135 | 22 | |
| Income Level†, n (%) | | | | |
| Below $40k | 38 (26.0) | 28 (23.1) | 10 (40.0) | 0.1957 |
| $40k-100k | 42 (28.8) | 37 (30.6) | 5 (20.0) | |
| 100k and above | 34 (23.3) | 27 (22.3) | 7 (28.0) | |
| Information not provided by Participant | 32 (21.9) | 29 (24.0) | 3 (12.0) | |
| Data not yet availableᶲ | 159 | 137 | 22 | |
| Health Insurance†, n (%) | | | | |
| Insured | 143 (97.9) | 120 (99.2) | 23 (92.0) | 0.0760 |
| Uninsured | 3 (2.1) | 1 (0.8) | 2 (8.0) | |
| Data not yet availableᶲ | 159 | 137 | 22 | |
| Marital Status†, n (%) | | | | |
| Not married | 38 (26.0) | 30 (24.8) | 8 (32.0) | 0.0858 |
| Married | 107 (73.2) | 91 (75.2) | 16 (64.0) | |
| Information not provided by Participant | 1 (0.8) | 0 (0.0) | 1 (4.0) | |
| Data not yet availableᶲ | 159 | 137 | 22 | |
| Family History of Pancreatic Cancer†, n (%) | | | | |
| No | 87 (67.4) | 75 (69.4) | 12 (57.1) | 0.0779 |
| Yes | 16 (12.4) | 15 (13.9) | 1 (4.8) | |
| Participant does not know | 26 (20.2) | 18 (16.7) | 8 (38.1) | |
| Data not yet availableᶲ | 176 | 150 | 26 | |
| Distress‡, n (%) | | | | |
| No | 36 (12.4) | 35 (14.3) | 1 (2.2) | 0.025 |
| Yes | 255 (87.6) | 210 (85.7) | 45 (97.8) | |
| Data not yet availableᶲ | 14 | 13 | 1 | |
| Depression‡, n (%) | | | | |
| No | 189 (64.9) | 157 (64.1) | 32 (69.6) | 0.5707 |
| Mild depression | 43 (14.8) | 39 (15.9) | 4 (8.7) | |
| Moderate depression | 40 (13.8) | 34 (13.9) | 6 (13.0) | |
| Severe depression | 19 (6.5) | 15 (6.1) | 4 (8.7) | |
| Data not yet availableᶲ | 14 | 13 | 1 | |
| Smoking status†‡, n (%) | | | | |
| No | 129 (44.3) | 109 (44.3) | 20 (44.4) | 0.4935 |
| Former smoker | 127 (43.6) | 105 (42.7) | 22 (48.9) | |
| Current smoker | 35 (12.1) | 32 (13.0) | 3 (6.7) | |
| Data not yet availableᶲ | 14 | 12 | 2 | |
| Marijuana status†, n (%) | | | | |
| No | 97 (71.9) | 76 (68.5) | 21 (87.5) | 0.2575 |
| Former user | 25 (18.5) | 23 (20.7) | 2 (8.3) | |
| Current user | 13 (9.6) | 12 (10.8) | 1 (4.2) | |
| Data not yet availableᶲ | 170 | 147 | 23 | |
| Abdominal Pain¶, n (%) | | | | |
| No | 78 (38.8) | 72 (41.4) | 6 (22.2) | 0.1512 |
| Yes | 100 (49.8) | 83 (47.7) | 17 (63.0) | |
| Information unavailble in EMR | 23 (11.4) | 19 (10.9) | 4 (14.8) | |
| Data not yet availableᶲ | 104 | 84 | 20 | |

| | | | | |
|---|---|---|---|---|
| Fatigue[¶], n (%) | | | | |
| No | 148 (52.5) | 125 (52.3) | 23 (53.5) | 0.1754 |
| Yes | 98 (34.7) | 80 (33.5) | 18 (41.9) | |
| Information unavailble in EMR | 36 (12.8) | 34 (14.2) | 2 (4.6) | |
| Data not yet available[ϕ] | 23 | 19 | 4 | |
| GI Bleeding[¶], n (%) | | | | |
| No | 217 (77.0) | 181 (75.7) | 36 (83.7) | 0.0269 |
| Yes | 7 (2.5) | 4 (1.7) | 3 (7.0) | |
| Information unavailble in EMR | 58 (20.5) | 54 (22.6) | 4 (9.3) | |
| Data not yet available[ϕ] | 23 | 19 | 4 | |
| Jaundice[¶], n (%) | | | | |
| No | 178 (62.9) | 152 (63.3) | 26 (60.5) | 0.1893 |
| Yes | 66 (23.3) | 52 (21.7) | 14 (32.5) | |
| Information unavailble in EMR | 39 (13.8) | 36 (15.0) | 3 (7.0) | |
| Data not yet available[ϕ] | 22 | 18 | 4 | |
| Weight Loss More Than 5%[¶], n (%) | | | | |
| No | 133 (47.4) | 115 (48.3) | 18 (41.9) | 0.7094 |
| Yes | 115 (40.9) | 95 (39.9) | 20 (46.5) | |
| Information unavailble in EMR | 33 (11.7) | 28 (11.8) | 5 (11.6) | |
| Data not yet available[ϕ] | 24 | 20 | 4 | |
| Charlsons Comorbidity Index, n (%) | | | | |
| 0 | 164 (57.7) | 142 (58.9) | 22 (51.2) | 0.1975 |
| <=2 | 101 (35.6) | 81 (33.6) | 20 (46.5) | |
| >=3 | 19 (6.7) | 18 (7.5) | 1 (2.3) | |
| Data not yet available[ϕ] | 21 | 17 | 4 | |
| Personal History of Diabetes[†¶], n (%) | | | | |
| No | 195 (68.4) | 167 (69.0) | 28 (65.1) | 0.6129 |
| Yes | 90 (31.6) | 75 (31.0) | 15 (34.9) | |
| Data not yet available[ϕ] | 20 | 16 | 4 | |
| Personal History of Pancreatitis[†¶], n (%) | | | | |
| No | 180 (79.6) | 158 (81.0) | 22 (71.0) | 0.1964 |
| Yes | 46 (20.4) | 37 (19.0) | 9 (29.0) | |
| Data not yet available[ϕ] | 79 | 63 | 16 | |
| Cachexia[‡¶], n (%) | | | | |
| refractory cachexia | 10 (3.8) | 5 (2.3) | 5 (11.6) | 0.0039 |
| cachexia | 76 (29.1) | 59 (27.0) | 17 (39.5) | |
| pre-cachexia | 26 (10.0) | 25 (11.5) | 1 (2.4) | |
| non cachectic | 149 (57.1) | 129 (59.2) | 20 (46.5) | |
| missing | 44 | 40 | 4 | |
| Body Mass Index (kg/m2)[¶]<br>n, mean (SD) | 281, 27 (5.5) | 235, 27 (5.5) | 46, 26 (5.5) | 0.1791 |
| Waist Circumference,[¶]<br>n, mean (SD) | 231, 40 (12.7) | 191, 40 (12.7) | 40, 40 (12.7) | 0.9476 |
| Histology[¶], n (%) | | | | |
| Pancreatic Ductal Adenocarcinoma (PDAC) | 183 (61.4) | 154 (61.1) | 29 (63.0) | 0.0064 |
| Pancreatic Neuroendocrine Tumor (PNET) | 35 (11.7) | 33 (13.1) | 2 (4.4) | |
| Intraductal Papillary Mucinous Neoplasm (IPMN) | 35 (11.7) | 33 (13.1) | 2 (4.4) | |
| Mucinous Cystic Neoplasm (MCN) | 6 (2.0) | 6 (2.4) | 0 (0.0) | |
| Other[§] | 39 (13.1) | 26 (10.3) | 13 (28.2) | |
| Data not yet available[ϕ] | 7 | 6 | 1 | |
| Surgical Resection Attempted[¶], n (%) | | | | |
| No | 146 (47.9) | 116 (45.0) | 30 (63.8) | |
| Yes | 159 (52.1) | 142 (55.0) | 17 (36.2) | |
| Location of Tumor[¶], n (%) | | | | |
| Body | 16 (14.0) | 16 (14.0) | 0 (0.0) | 0.5527 |
| Diffuse | 17 (14.9) | 15 (13.2) | 2 (20.0) | |
| Head | 65 (57.0) | 60 (52.6) | 5 (50.0) | |
| Tail | 13 (11.4) | 12 (10.5) | 1 (10.0) | |
| Other | 13 (11.4) | 11 (9.7) | 2 (20.0) | |
| Data not yet available[ϕ] | 181 | 144 | 37 | |

| | | | | |
|---|---|---|---|---|
| Stage¶, n (%) | | | | |
| Stage 0 | 22 (14.1) | 20 (14.8) | 2 (13.3) | <.0001 |
| Stage I/II | 87 (55.8) | 85 (63.0) | 2 (13.3) | |
| Stage III/IV | 41 (26.3) | 30 (22.2) | 11 (73.3) | |
| Data not yet availableᶲ | 155 | 123 | 32 | |
| Grade Exocrine Pancreatic Tumor¶¥, n (%) | | | | |
| Well diffferentiated | 7 (8.0) | 7 (8.7) | 0 (0.0) | 0.0804 |
| Moderately differentiated | 29 (32.9) | 29 (36.3) | 0 (0.0) | |
| Poorly differentiated | 21 (23.9) | 18 (22.5) | 3 (37.5) | |
| Grade undetermined | 31 (35.2) | 26 (32.5) | 5 (62.5) | |
| Data not yet availableᶲ | 136 | 113 | 23 | |
| Grade IPMN¶, n (%) | | | | |
| Low grade | 12 (34.3) | 12 (36.4) | 0 (0.0) | 0.6975 |
| Borderline | 2 (5.7) | 2 (6.1) | 0 (0.0) | |
| Carcinoma-in-situ | 5 (14.3) | 5 (15.1) | 0 (0.0) | |
| Invasive carcinoma | 1 (2.9) | 1 (3.0) | 0 (0.0) | |
| Unknown | 15 (42.9) | 13 (39.4) | 2 (100.0) | |
| Positive Lymph Nodes¶, n (%) | | | | |
| No | 110 (94.8) | 100 (95.2) | 10 (90.9) | 0.4576 |
| Yes | 6 (5.2) | 5 (4.8) | 1 (9.1) | |
| Data not yet availableᶲ | 189 | 153 | 36 | |

†Data available from partially-or fully-completed baseline questionnaires at time of analysis.
ᶲData not yet available. Additional data will be included in future analyses when entered into the DatStat system.
‡Data available from the health screen questionnaire at time of analysis.
¶Data available from case report forms (CRFs) at time of analysis.
§The 'Other' category includes benign and malignant tumors of pancreatic, liver and bile duct, renal, adrenal gland, lymph node, and unclassified origin.
¥The grade for exocrine tumor has been restricted to PDAC, IPMN, and MCN (n=224).
